# Effect of protocolized MAGNesium replacement in critically ill patients On mortaLIty and Atrial fibrillation: protocol for the MAGNOLIA randomized controlled trial

**DOI:** 10.64898/2026.11.03.26366752

**Authors:** Christopher J Yarnell, Selasi Bruce-Kemevor, Karen Burns, Lisa Burry, Joshua Craig, Shannon M Fernando, Rob Fowler, Keith Gunaratne, Jan Haskings, Francois Lamontagne, Kuan Liu, Varuna Prakash, Stephanie Sibley, Karim Soliman, George Tomlinson, Louis Wei, Thomas Bodley, Canadian Critical Care Trials Group

**Affiliations:** University of Toronto; Scarborough Health Network Research Institute, Scarborough, Toronto, Canada; Department of Critical Care Medicine, Unity Health; and Interdepartmental Division of Critical Care Medicine, University of Toronto, Toronto, Canada; Departments of Pharmacy, Medicine, Sinai Health Research, Sinai Health; and Leslie Dan Faculty of Pharmacy and Interdepartmental Division of Critical Care, University of Toronto, Toronto, Canada; Lakeridge Health Department of Critical Care Medicine, Oshawa, Canada; Department of Critical Care Medicine, Queen's University, Kingston, Canada; Sunnybrook Health Sciences Centre, Toronto, Canada; Department of Medicine and Interdepartmental Division of Critical Care Medicine, University of Toronto, Toronto, Canada; Department of Critical Care Medicine, Oak Valley Health, Markham, Canada; Patient Family Advisor Committee, Scarborough Health Network, Toronto, Ontario, Canada; Department of Medicine, Faculty of Medicine, University of Sherbrooke; Centre Hospitalier Universitaire de Sherbrooke, Sherbrooke, Quebec, Canada; Institute of Health Policy, Management, and Evaluation, University of Toronto, Toronto, Canada; Department of Critical Care Medicine, Queen's University, Kingston, Canada; University Health Network, Toronto, Canada; University of Toronto, Toronto, Canada; Department of Pharmacy and Critical Care Medicine, Scarborough Health Network, Toronto, Canada; Department of Critical Care Medicine and Research Institute, Scarborough Health Network, Toronto, Canada; Department of Medicine and Interdepartmental Division of Critical Care Medicine, University of Toronto, Toronto, Canada

**Keywords:** *Magnesium*, *atrial fibrillation*, *critical illness*, *protocols*, *routine care*, *digital health*

## Abstract

**Background:** Measurement of serum magnesium levels, and administration of supplemental magnesium when levels are below target, is a common element of routine care for critically ill patients. However, targets for replacement vary, and the targets that lead to the best outcomes are unknown.

**Methods:** Multi-center open-label parallel group randomized controlled superiority trial of adult critically ill patients receiving protocolized magnesium replacement, comparing a higher target (>0.95mmol/L) to a lower target (>0.7mmol/L). The trial will be embedded into the electronic health record (EHR) at 6 hospitals across 2 health networks in Ontario, Canada, with a shared EHR. We will include patients aged 16 years or older who have ICU admission orders, including an order for the magnesium replacement protocol. Patients with pre-eclampsia, sustained ventricular tachycardia, or neuromuscular junction disease will be excluded. Because both targets lie within the standard of care, we will use an opt-out consent approach. Screening will be integrated with the EHR, such that when a patient meets inclusion criteria, a pop-up will appear to allow the ordering clinician to confirm eligibility or indicate the relevant exclusion. After the clinician confirms eligibility, patients will be randomly assigned to a magnesium target. The primary outcome will be an ordinal composite, evaluated at 30 days, composed of death and the number of days free of atrial fibrillation or flutter in ICU. Secondary outcomes will include ventricular arrhythmia and antiarrhythmics administered; receipt of vasopressors, ventilation, and new renal replacement therapy in ICU; lengths of ICU and hospital stay; hospital mortality at 60 and 90 days, magnesium levels, and magnesium supplementation. For analyses, we will use Bayesian regression with weakly skeptical priors and an intention-to-treat approach. Outcome ascertainment will occur within the EHR.

**Discussion:** This randomized controlled trial addresses an important uncertainty regarding routine care in the ICU with an EHR-embedded design. The innovative EHR-embedded design facilitates the large sample sizes and comprehensive, equitable recruitment needed for a trial evaluating a routine care intervention, and will lead to seamless integration with routine care upon trial completion.

**Trial registration:** ClinicalTrials.gov, NCT07173855. Registered 12 September 2025, https://clinicaltrials.gov/study/NCT07173855.

## Administrative information

Note: the numbers in curly brackets in this protocol refer to SPIRIT checklist item numbers. The order of the items has been modified to group similar items (see http://www.equator-network.org/reporting-guidelines/spirit-2013-statement-defining-standard-protocol-items-for-clinical-trials/).

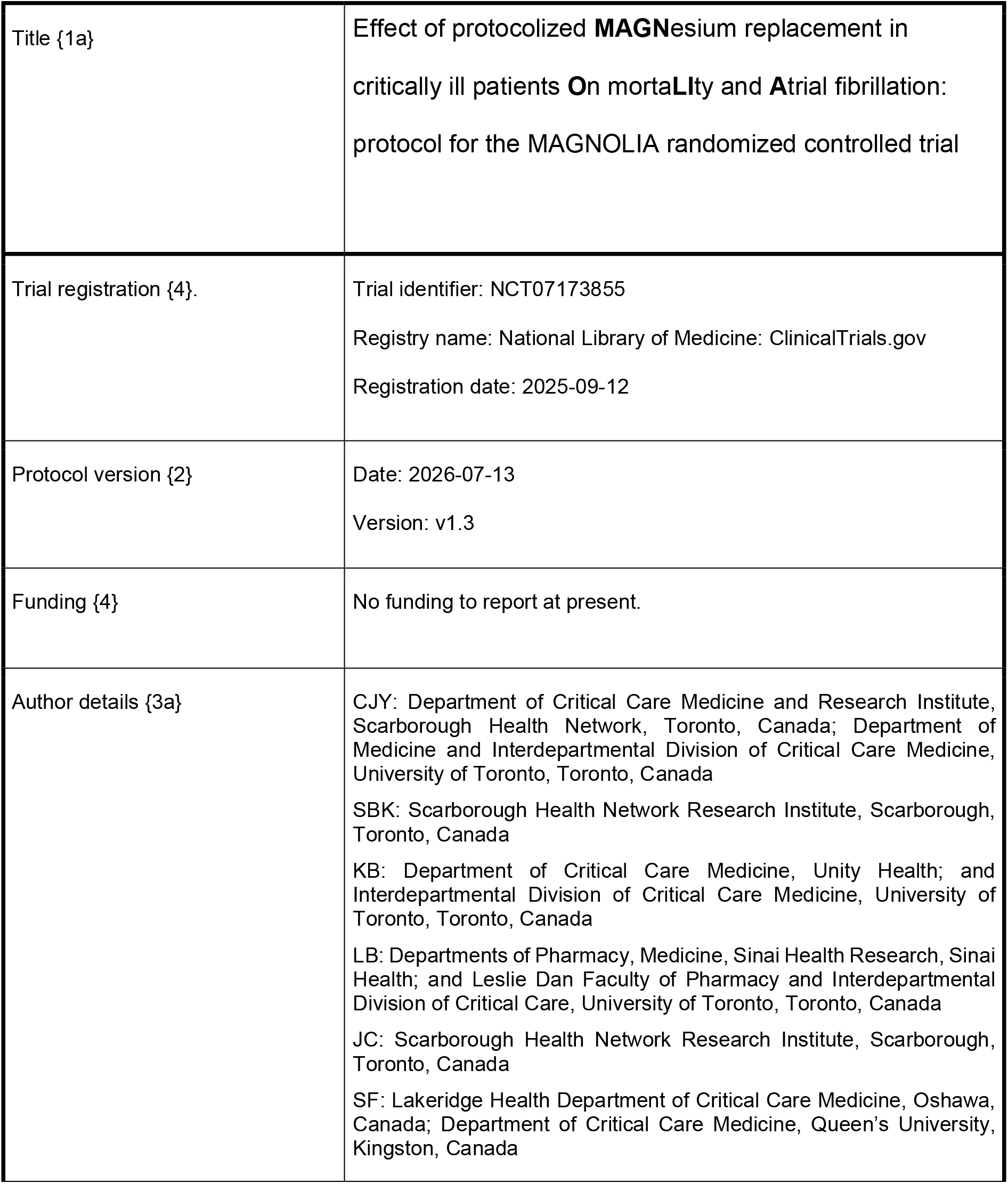
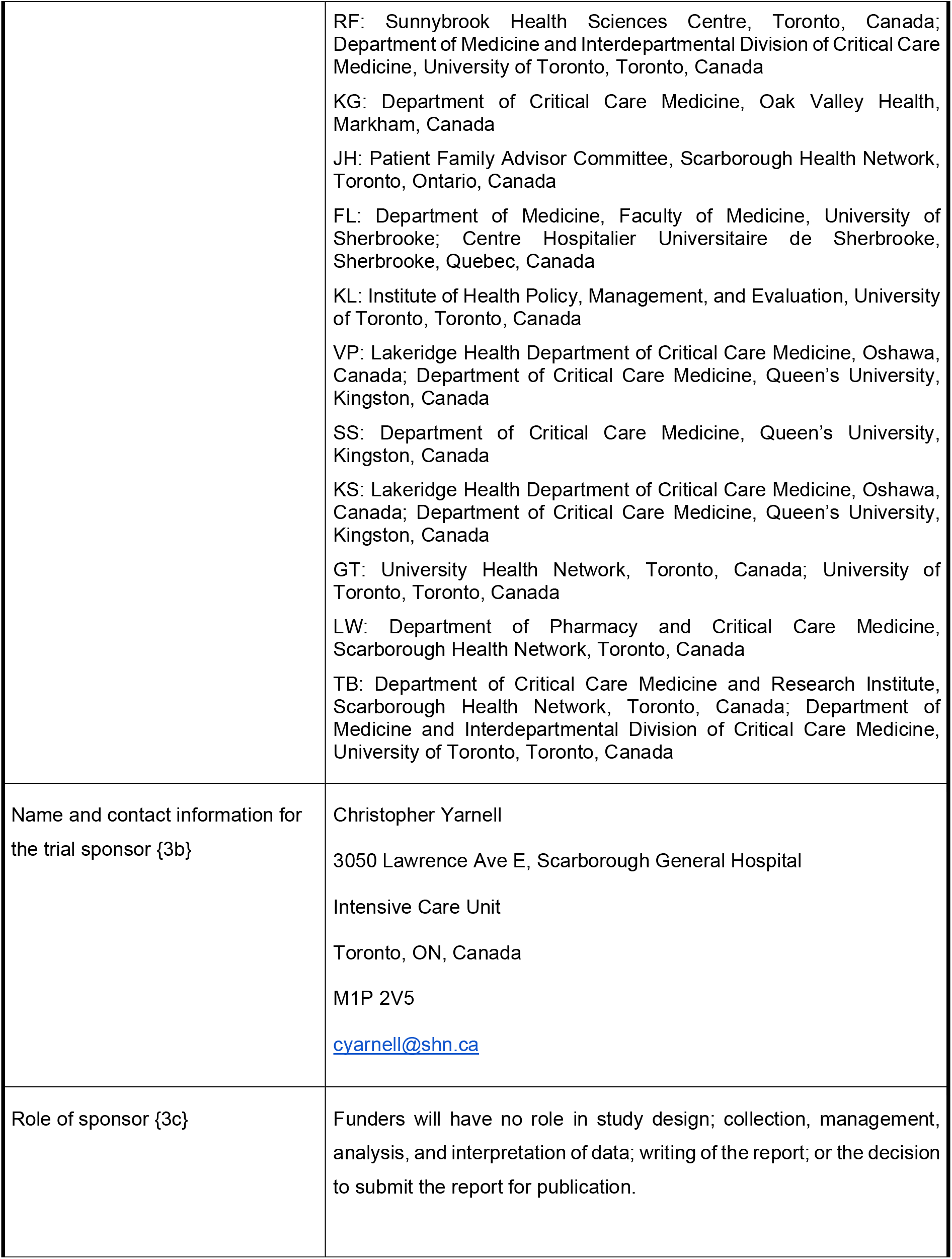
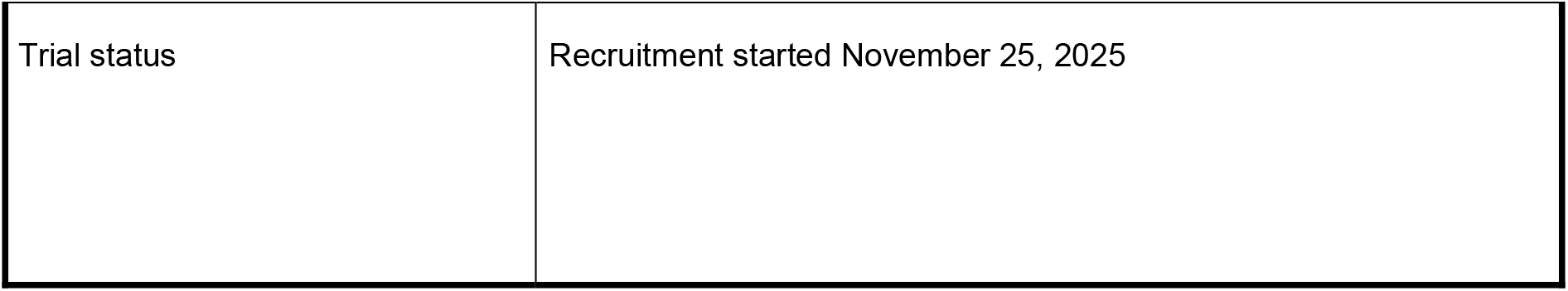

## Amendments

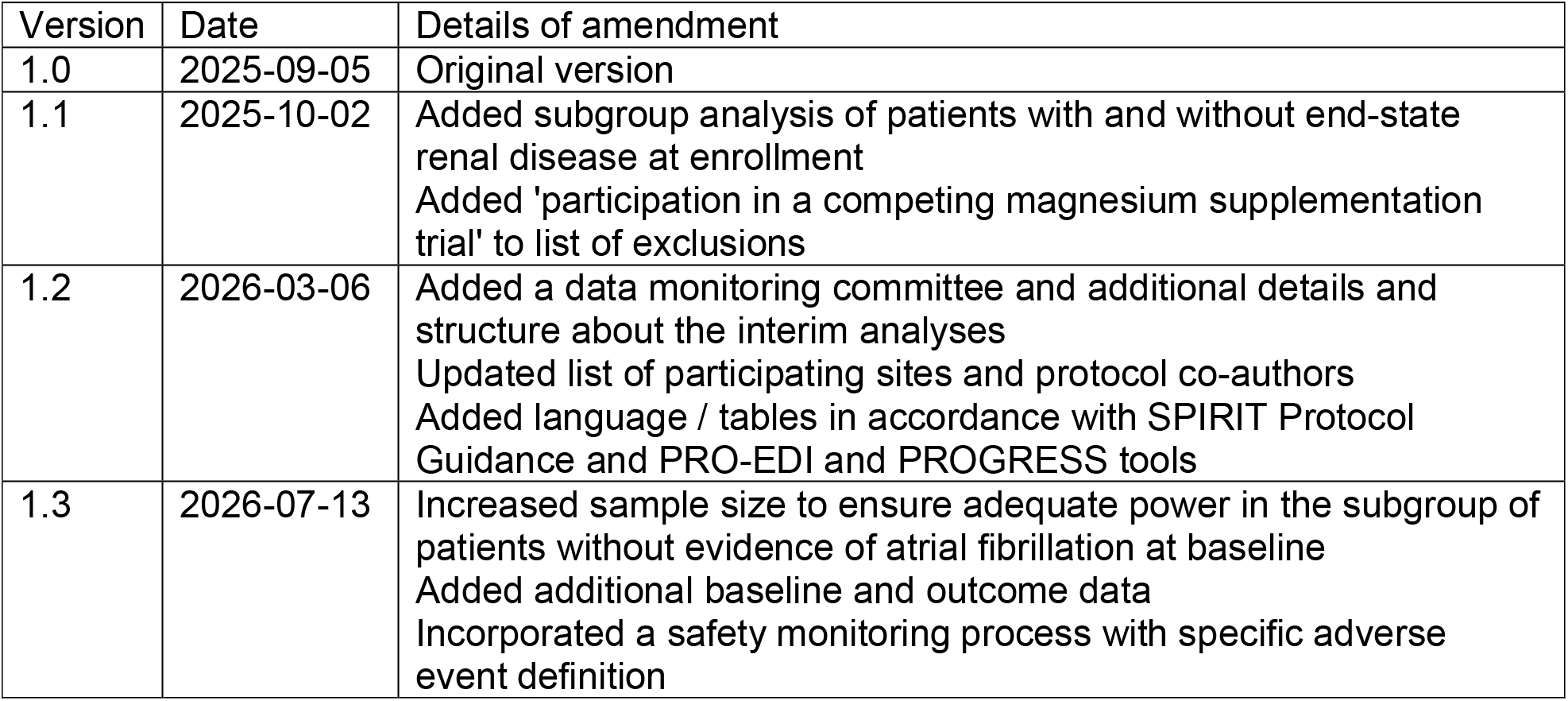

| Version | Date | Details of amendment |
|---|---|---|
| 1.0 | 2025-09-05 | Original version |
| 1.1 | 2025-10-02 | Added subgroup analysis of patients with and without end-state renal disease at enrollment<br>Added 'participation in a competing magnesium supplementation trial' to list of exclusions |
| 1.2 | 2026-03-06 | Added a data monitoring committee and additional details and structure about the interim analyses<br>Updated list of participating sites and protocol co-authors<br>Added language / tables in accordance with SPIRIT Protocol Guidance and PRO-EDI and PROGRESS tools |
| 1.3 | 2026-07-13 | Increased sample size to ensure adequate power in the subgroup of patients without evidence of atrial fibrillation at baseline<br>Added additional baseline and outcome data<br>Incorporated a safety monitoring process with specific adverse event definition |

## Introduction

### Background and rationale {9a}

Magnesium is essential for cellular function, with important roles in regulating cardiac rhythm, vascular tone, and the nervous system.(1) Low magnesium levels are common in critically ill patients, and clinical consequences include cardiac dysrhythmias such as atrial fibrillation and flutter.(2) New-onset atrial fibrillation has been associated with increased hospital mortality and costs among critically ill patients.(3–5) Meta-analyses of randomized trials show that intravenous magnesium may prevent atrial fibrillation after cardiac surgery(6–8), however, in non-cardiac critically ill patients, the effects are less certain.(9,10) No studies have evaluated the practice of protocolized magnesium administration based on serum levels (replacement) in critically ill patients.

Protocolized magnesium replacement is common. In an international survey of 283 mixed ICUs, 66% used magnesium replacement protocols.(11) The most common threshold for replacement was a serum level less than the lower limit of normal, however, some protocols recommend replacement while levels are still within the normal range for some or all patients.(11–13) Intravenous magnesium sulfate is the most common replacement agent(14,15), although oral options exist.(16) The optimal threshold serum level for routine protocolized magnesium replacement is unknown.

Higher target magnesium levels (eg. 0.95 mmol/L) increase the likelihood of replacement and may reduce the incidence of atrial fibrillation in non-cardiac-surgical critically ill patients. Such thresholds approximate routine administration of magnesium, because 86% of critically ill patients have a magnesium level of 0.95 mmol/L or less at ICU admission.(2) Observational studies conflict on whether magnesium supplementation reduces tachyarrhythmia.(13,17) A meta-analysis of 4 randomized controlled trials (RCTs), representing 4,654 patients, showed an odds ratio (OR) of 0.72 (95% confidence interval 0.48 to 1.09) for reduction of new-onset atrial fibrillation with magnesium administration compared to usual care.(9) However, most patients were from a trial enrolling patients with acute myocardial infarction in a coronary care unit(18) and a trial enrolling patients with pre-hospital acute stroke.(19) The applicability of these findings to contemporary patients with sepsis, respiratory failure, or other critical illness is unclear.

Higher targets and the resulting magnesium supplementation may also reduce mortality. A meta-analysis of 3 RCTs enrolling critically ill patients (n = 235) found a relative risk for mortality of 0.54 (95% confidence interval 0.30 to 0.96) with magnesium supplementation compared to no supplementation (very low certainty of evidence).(10) A recent pseudo-randomized observational study showed an association between protocolized magnesium replacement at a threshold of 0.95 mmol/L and reduced incidence of a composite outcome of death, ventricular and supraventricular arrhythmia, and atrial fibrillation or flutter over the next 24 hours (OR 0.87, 95% credible interval 0.74 to 1.01, probability of OR < 1 = 0.97)(13), while another study using a similar methodology found no benefit with supplementation (17). Randomized evidence to assess the benefits of a higher threshold is needed.

The alternative to a higher target is a lower target (eg. replacement when magnesium is 0.7mmol/L or less). Lower targets lead to fewer doses of electrolytes, which reduces fluid overload(20,21), costs(21), nursing workload(22), and environmental waste.(20) Avoiding unnecessary fluid administration is important, because positive cumulative fluid balance is associated with mortality.(23) Furthermore, although magnesium administration is generally safe, rare cases exist linking severe iatrogenic hypermagnesemia to coma and cardiac arrest.(24,25)

Given these patient, environment, and cost considerations, it is crucial to investigate whether protocolized magnesium replacement aiming at a higher (0.95 mmol/L) or lower (0.7 mmol/L) target leads to lower incidence of mortality and atrial fibrillation in critically ill patients. To this end, we propose an RCT, described in this protocol according to the Standard Protocol Items: Recommendations for Interventional Trials guidelines.(26)

### Objectives {10}

To compare protocolized magnesium replacement targeting higher (magnesium > 0.95 mmol/L) and lower (magnesium > 0.7 mmol/L) levels in adult patients with critical illness, with respect to:

1. Burden of atrial fibrillation
2. Hospital mortality
3. Organ support (ventilator, vasopressor, and renal replacement therapy use)
4. Costs of magnesium replacement
5. Nursing workload
6. Carbon footprint.

### Trial design {12}

This is a pragmatic, comparative effectiveness, parallel group superiority trial.(27) We will compare magnesium replacement protocols targeting higher (magnesium > 0.95 mmol/L) and lower (magnesium > 0.7 mmol/L) magnesium levels in adult patients with critical illness. The allocation ratio will be 1:1. The trial will be embedded into the electronic health record (EHR) with respect to screening, enrollment, randomization, assignment, follow-up, and outcome ascertainment, in order to efficiently capture an unselected population of eligible critically ill patients and create seamless local implementation at the end of the trial (Figure 1).(20,28)

**Figure 1:**
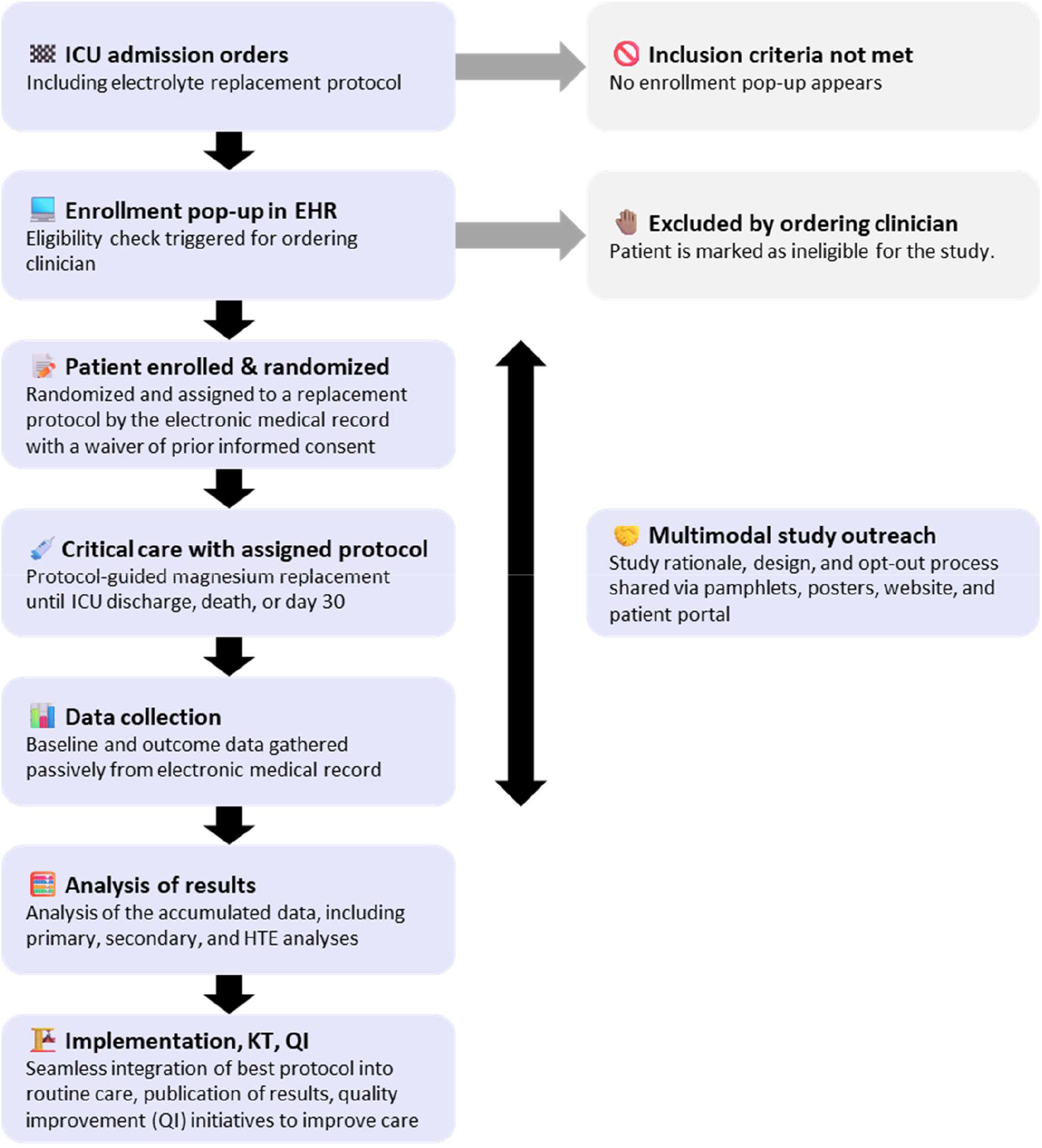
Study design overview. Caption: This figure shows an overview of the study design, starting from the moment of screening (top left), proceeding through the study (left side) to post-study activities (bottom left). The diagram also includes descriptions of ways in which patients would be ineligible for the trial (right, gray) and a description of the multimodal outreach strategy (right, blue). Note that patients can opt out at any time; if they opt out after study interventions are finished then we remove their data from the study. Mg = magnesium, EHR = electronic health record, QI = quality improvement, SDM = substitute decision maker, ICU = intensive care unit, HTE = heterogeneity of treatment effect.

### Patient and public involvement {11}

Trial design and conduct benefit from the advice of a patient-family partner (JH). We also consulted with the Scarborough Health Network Patient-Family Advisor Committee regarding the approach to consent (Appendix B).

## Methods: Participants, interventions and outcomes

### Study setting {13}

The study will take place in medical-surgical critical care units at 6 hospitals which use a shared EHR: Scarborough General Hospital, Scarborough Centenary Hospital, Scarborough Birchmount Hospital, Lakeridge Health Oshawa Hospital, the Lakeridge Health Ajax-Pickering Hospital, and the Lakeridge Health Bowmanville Hospital, all in Ontario, Canada. These hospitals serve a diverse urban and suburban area (Table 1).

**Table 1:**
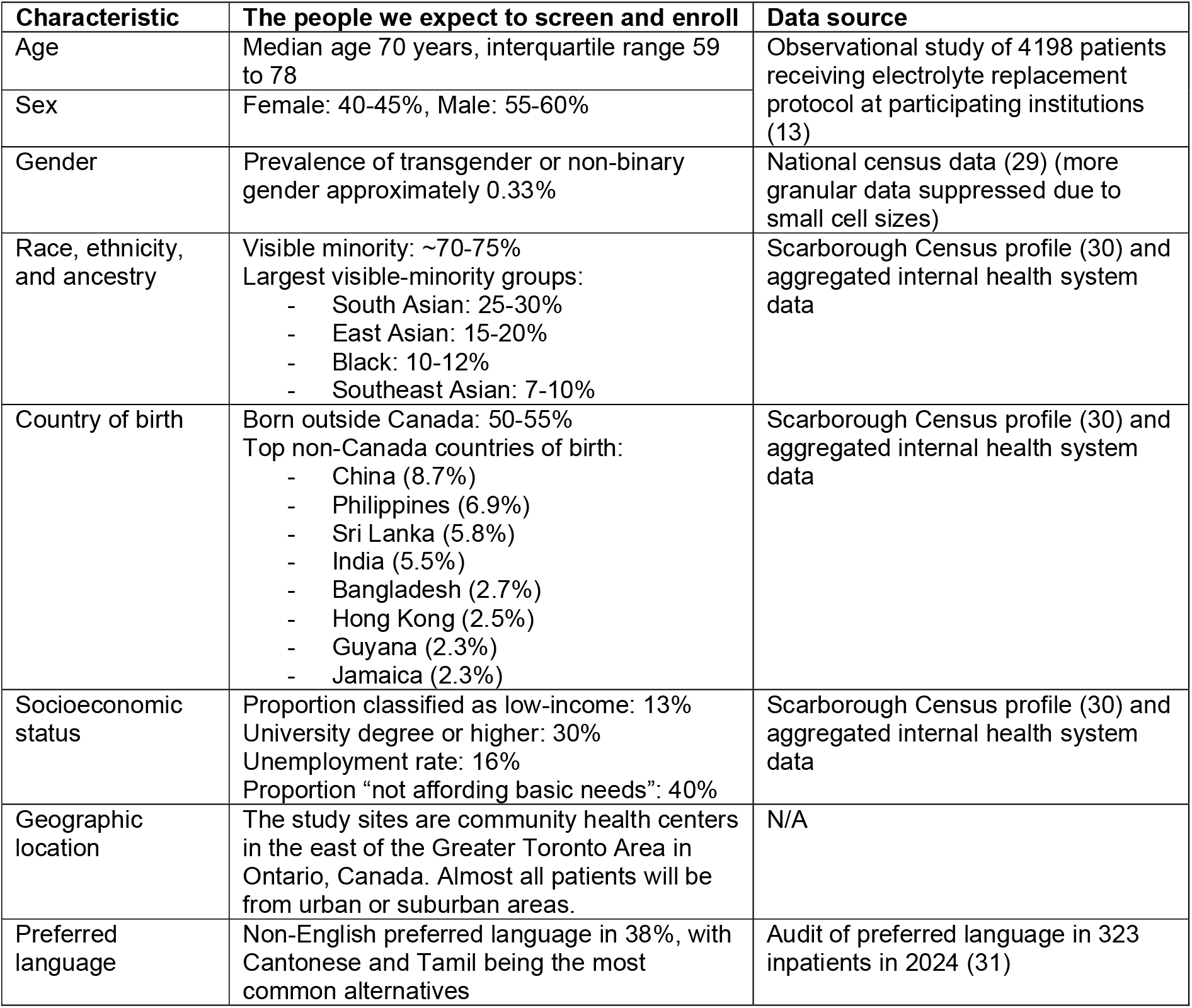
Non-clinical participant characteristics.

| Characteristic | The people we expect to screen and enroll | Data source |
|---|---|---|
| Age | Median age 70 years, interquartile range 59 to 78 | Observational study of 4198 patients receiving electrolyte replacement protocol at participating institutions (13) |
| Sex | Female: 40-45%, Male: 55-60% |  |
| Gender | Prevalence of transgender or non-binary gender approximately 0.33% | National census data (29) (more granular data suppressed due to small cell sizes) |
| Race, ethnicity, and ancestry | Visible minority: ~70-75%<br>Largest visible-minority groups:<br>- South Asian: 25-30%<br>- East Asian: 15-20%<br>- Black: 10-12%<br>- Southeast Asian: 7-10% | Scarborough Census profile (30) and aggregated internal health system data |
| Country of birth | Born outside Canada: 50-55%<br>Top non-Canada countries of birth:<br>- China (8.7%)<br>- Philippines (6.9%)<br>- Sri Lanka (5.8%)<br>- India (5.5%)<br>- Bangladesh (2.7%)<br>- Hong Kong (2.5%)<br>- Guyana (2.3%)<br>- Jamaica (2.3%) | Scarborough Census profile (30) and aggregated internal health system data |
| Socioeconomic status | Proportion classified as low-income: 13%<br>University degree or higher: 30%<br>Unemployment rate: 16%<br>Proportion “not affording basic needs”: 40% | Scarborough Census profile (30) and aggregated internal health system data |
| Geographic location | The study sites are community health centers in the east of the Greater Toronto Area in Ontario, Canada. Almost all patients will be from urban or suburban areas. | N/A |
| Preferred language | Non-English preferred language in 38%, with Cantonese and Tamil being the most common alternatives | Audit of preferred language in 323 inpatients in 2024 (31) |

### Eligibility criteria for participants {14a}

Inclusion criteria:

1. Adult (age 16 years or more)
2. Order placed for admission to a medical-surgical intensive care unit (ICU)
3. Order placed for the magnesium replacement protocol.

Exclusion criteria:

1. Current diagnosis of eclampsia or pre-eclampsia
2. Ongoing concern for sustained ventricular tachycardia
3. Active neuromuscular junction disease (eg. myasthenia gravis)
4. Prior enrollment in or withdrawal from the MAGNOLIA trial
5. Enrollment in a separate trial that prevents administration of the MAGNOLIA intervention (for example, a trial of magnesium supplementation vs no supplementation)

### Approach to consent {32a}

We will use a waiver of prior informed consent with the opportunity for patients to subsequently opt out of the study, on the basis of both intervention arms being minimal risk and within the standard of care, the time-sensitive nature of enrollment, and the impracticability of achieving informed consent on either an a priori or deferred basis for all patients admitted to the ICU.

### Rationale for opt-out consent

<u>The intervention arms are minimal risk and within the standard of care.</u> The higher threshold is currently used at the participating ICUs. The lower threshold is consistent with clinical practice at many other centers, both locally and internationally.(11)

<u>Enrollment is time-sensitive.</u> Electrolyte replacement protocols are ordered at the time of ICU admission. To best reflect their pragmatic, real-life use, enrollment into the trial needs to reflect this. The assignment also needs to be known at ICU admission, because 54% of admission magnesium values lie between 0.7 and 0.95 mmol/L at SHN and LH, similar to other data.(2,32,33)

<u>Informed consent is impracticable.</u> Obtaining informed a priori or deferred consent will be impracticable in this case, because of the volume of enrolments needed to ensure an unselected and scientifically applicable patient population. Electrolyte protocols are used in almost all critically ill patients, and therefore almost all critically ill patients need to be enrolled in the study.

<u>An opt-out approach is consistent with current practice.</u> Magnesium replacement protocols are a standard element of routine care, and clinicians do not seek informed consent regarding these protocols. Protocols are generally implemented at the hospital or unit level, and in many centers the thresholds are not adjusted for individual patients (for example, at participating centres). There are precedents for opt-out or waived consent in critical care comparative effectiveness research.(20,28,34)

<u>Patients support opt-out consent in this context</u>, provided that participants are made aware of their enrollment in the trial.(35–38)(summary of consultation with SHN Patient-Family Advisor Committee in Supplement) Opt-out consent may also improve the representation of non-white patients in research studies.(39)

### Operationalizing opt-out consent

We will ensure that all patients who desire to opt out will have the opportunity to do so by educating bedside staff about the study, placing posters in the every patient room and throughout common areas in the study ICUs, distributing a pamphlet about the study to each admitted patient or their family, developing a study website with QR code available on the pamphlets and posters (example materials in Appendix D), and posting study information on the online patient portal. All study materials will be translated into common non-English languages at participating sites.(31)

Patients and SDMs will be able to contact the research team to opt out of the study by: 1) mentioning this to the clinical team, 2) emailing the study email address, or 3) calling the research team phone number. The research team will then reach out to the patient or SDM to clarify their rationale for opting out and complete the opt-out process. More details are available in Appendix A. If a patient opts out while still in the ICU, their magnesium protocol will revert to the higher threshold (which is the pre-study protocol at participating sites).

To further support the principle of respect for participants, the EHR-embedded nature of the interventions will help ensure robust implementation and uptake of trial findings.(40)

### Additional consent provisions for collection and use of participant data and biological specimens {32b}

We do not plan to seek consent for additional use of participant data or biological specimens.

## Interventions

After clinicians confirm eligibility, patients will be randomly assigned to a magnesium target. The intervention protocols are specified in Table 2.

**Table 2:**
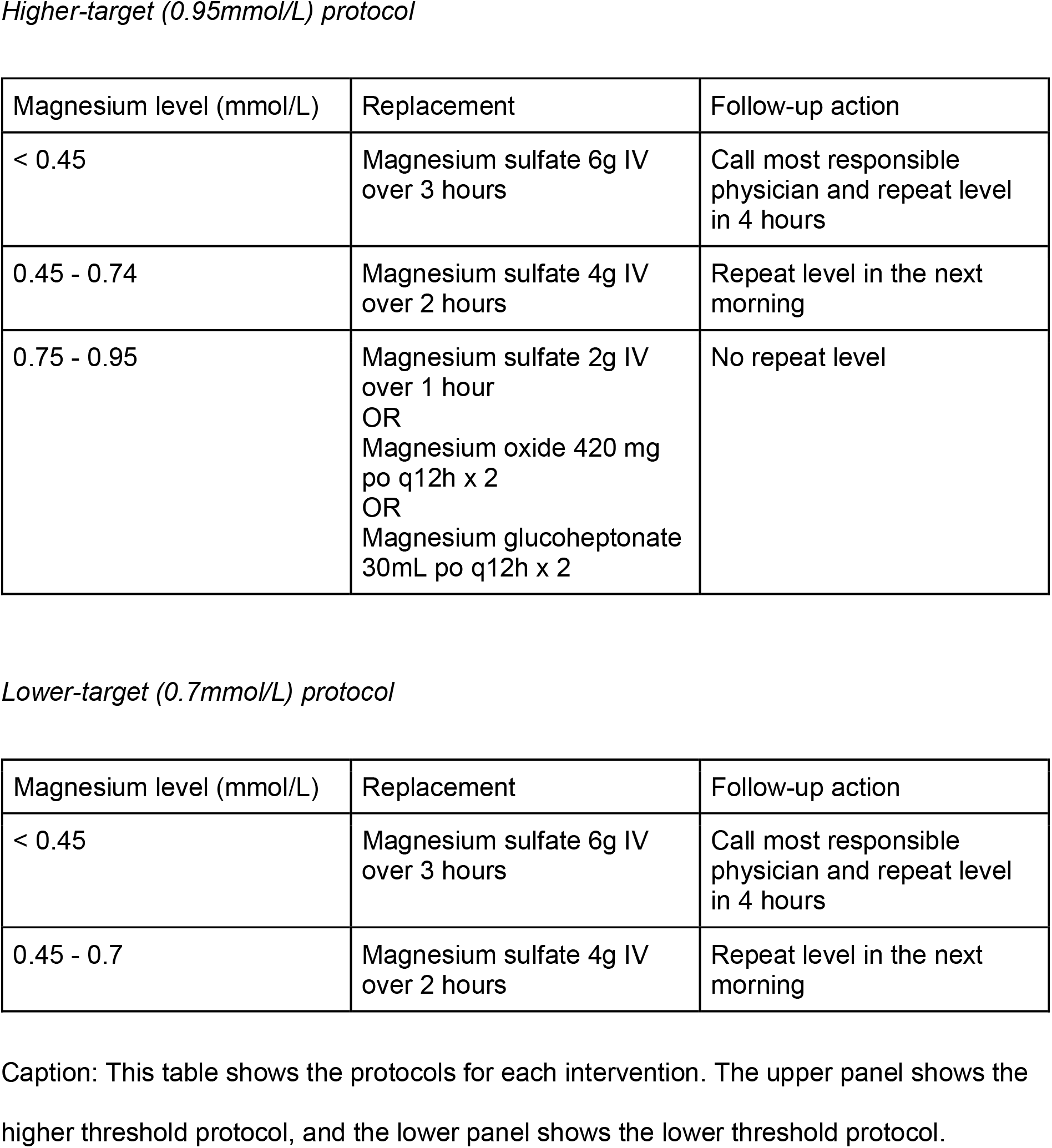
Intervention protocols.

### Explanation for the choice of comparators {9b}

The trial will compare magnesium replacement protocols with higher (>0.95 mmol/L) and lower (>0.7 mmol/L) targets. The rationale is that these options span but remain within the region of equipoise suggested by usual care across different hospitals and ICUs. Magnesium replacement is a Health Canada-approved use of magnesium.(41)

### Intervention description {15a}

The magnesium replacement protocol is ordered by the admitting intensivist. The EHR guides the bedside nurse to administer magnesium when serum magnesium levels are below certain thresholds. The thresholds will be determined by the assigned intervention. Interventions will be maintained during the ICU admission and any ICU readmissions for up to 30 days following study enrollment. After 30 days, the electrolyte protocol will revert to the higher threshold.

### Criteria for discontinuing or modifying allocated interventions {15b}

The electrolyte protocol will be discontinued at the time of discharge from the ICU. In clinically stable patients, the most responsible physician may elect to discontinue the protocol.

### Strategies to improve adherence to interventions {15c}

We will encourage adherence to the protocol through education of bedside nursing and physician staff via emails, weekly huddles, and material on the ICU research bulletin board. We will audit protocol deviations and provide feedback as described below.

### Relevant concomitant care permitted or prohibited during the trial {15d}

Physician-ordered additional magnesium sulfate (or oral preparations) will not be prohibited in either arm. In cases where physicians feel that more magnesium than that offered by protocol is needed, we will encourage them to order this in addition to the protocol, as opposed to discontinuing the protocol.

### Provisions for post-trial care {34}

We do not anticipate the need for any care provisions specific to the trial after the intervention period is finished.

## Baseline variables and study outcomes

### Baseline variables

For each enrolled participant, we will gather baseline data from the EHR. Baseline will be defined as the time of enrollment in the chart. These data will be used for subgroup and heterogeneity of treatment analyses (Table 3). The statistical details of these analyses will be fully elaborated in a future statistical analysis plan, but the main goal will be to identify subgroups of patients, using baseline characteristics, where the effect of higher or lower targets may differ from the overall average effect.

**Table 3:** Baseline variables.

| Type | Variable |
|---|---|
| Admission details | Hospital<br>Month and year of hospital admission<br>Department<br>Time of day<br>Day of week |
| Demographics and social determinants of health(42) | Age<br>Sex<br>Preferred language |
| Diagnoses | Primary diagnosis<br>Comorbidities |
| Laboratory values (most recent) | Hemoglobin<br>White blood cell count<br>Platelet count<br>Sodium<br>Potassium<br>Chloride<br>Bicarbonate<br>Creatinine<br>Magnesium<br>Calcium<br>Phosphate<br>Albumin<br>Alanine aminotransferase<br>Alkaline phosphatase<br>Bilirubin<br>Troponin<br>Lactate<br>Venous and arterial blood gas |
| Clinical data (most recent) | Glasgow Coma Scale<br>Respiratory rate<br>Oxygen device<br>Fraction of inspired oxygen<br>Oxygen flow (L/minute)<br>Work of breathing<br>Ventilator settings if ventilated (PEEP, PC/PS, Vt)<br>Heart rate<br>Cardiac rhythm<br>Blood pressure |
| Medications in last 24 hours | Norepinephrine<br>Vasopressin<br>Epinephrine |
|  | Milrinone<br>Dobutamine<br>Phenylephrine<br>Proton pump inihibitors<br>Diuretics (furosemide, metolazone, amiloride, spironolactone) |
| Echocardiogram data | Date of study relative to ICU admission<br>Left ventricular ejection fraction<br>Left atrial size<br>Tricuspid, aortic, or mitral stenosis (severe)<br>Tricuspid, aortic, or mitral regurgitation (severe) |

### Outcomes {16}

#### Primary outcome

The primary outcome will be a composite of hospital mortality and days free of atrial fibrillation or flutter in the ICU up to day 30 (Table 4). Among those who survive to 30 days, the outcome will be the number of days in the ICU where atrial fibrillation or flutter was documented in the electronic health record subtracted from 30. Hospital mortality will be scored as -1 (i.e., worse than 30 days of atrial fibrillation or flutter). For example, if a patient survives to day 30 and had atrial fibrillation charted on 4 days while in ICU, they would be recorded to have 26 days free of atrial fibrillation and flutter in the ICU. This outcome integrates both mortality and atrial fibrillation, which are the two clinical outcomes where magnesium has shown potential benefit. Tracking days with atrial fibrillation in ICU provides some insight into the burden of atrial fibrillation, which has been correlated with strokes and other adverse outcomes.(43)

**Table 4:** Outcome variables.

| Type | Variable |
|---|---|
| Primary | Composite combining hospital mortality at day 30 and days free of atrial fibrillation or flutter in ICU at day 30 |
| Secondary: evaluated at 30 days | Days free of ICU<br>Organ-support-free days<br>Days free of invasive ventilation<br>Days free of vasopressors<br>Days free of renal replacement therapy<br>Sustained ventricular tachycardia or ventricular fibrillation in ICU<br>Intravenous antiarrhythmics administered<br>Magnesium and potassium levels while in ICU<br>Magnesium administrations while in ICU (number, route, dose)<br>Daily fluid balance<br>Electrical cardioversion<br>Stroke |
| Secondary: DOOR, evaluated at 30 days | DOOR 1: Death, ventricular arrhythmia, atrial fibrillation or flutter, none of the above<br>DOOR 2: Discharge home, discharge to location other than home, ongoing hospital admission, persistent organ dysfunction in ICU, death. |
| Secondary: 60 or 90 days | Hospital mortality at day 60<br>Hospital mortality at day 90<br>Hospital length of stay at day 90<br>Discharge destination at day 90 |
| Additional outcomes | Nursing workload<br>Costs of magnesium administration<br>Carbon footprint of magnesium administration |

Caption: DOOR = desirability of outcome ranking, ICU = intensive care unit.

**Table 5:** Participant timeline {18}.

|  | **STUDY PERIOD** |  |  |
|---|---|---|---|
|  | Enrolment | Intervention | Study termination |
| **TIMEPOINT (day)** | ICU admission (time 0) | From time 0 to the earliest of death, ICU discharge, provider discontinuation of electrolyte protocol, or day 30 | Day 31+ |
| Eligibility screen | X |  |  |
| Admitting physician confirms eligibility | X |  |  |
| Randomization | X |  |  |
| Assignment | X |  |  |
| Bedside ICU nurse follows assigned protocol |  | X |  |
| Study material distributed to bedside |  | X |  |
| Outcome ascertainment |  |  | X |
| Opt-out possible |  | X | X |

#### Secondary outcomes

Secondary outcomes are hospital mortality at 30, 60, and 90 days; days with atrial fibrillation in the ICU up to day 30; days free of ICU up to day 30; organ-support-free days up to day 30 (with organ support defined as invasive ventilation, vasopressors, or new renal replacement therapy)(44); days free of invasive ventilation, days free of vasopressors, and days free of renal replacement therapy up to day 30; hospital length of stay and discharge destination up to 90 days; documented sustained ventricular tachycardia or ventricular fibrillation in ICU up to day 30; intravenous antiarrhythmics administered up to day 30 (amiodarone, metoprolol, esmolol, diltiazem, procainamide, lidocaine, flecainide, adenosine, digoxin); magnesium and potassium levels while in ICU up to day 30; number, route (intravenous vs enteral), and dose of magnesium administrations up to day 30; daily fluid balance while in ICU up to day 30.

We will also calculate two composite desirability of outcome ranking (DOOR) secondary outcomes evaluated at 30 days.(45) The first will be an ordinal composite of death, ventricular arrhythmia, atrial fibrillation or flutter, or none of the above. The second will be an ordinal composite of discharge to home, discharge to a location other than home, ongoing hospital admission, persistent organ dysfunction (vasopressor use, new renal replacement therapy, or invasive ventilation), or death.(44)

#### Additional outcomes

Additional outcomes are costs, nursing workload, and carbon footprint. We will calculate costs by obtaining the cost of each magnesium formulation and associated materials (vials, syringes, minibags, tubing), then multiplying the observed number of administrations by those costs. We will estimate nursing and pharmacy workload by observing a random sample of 20 magnesium administrations, then multiplying the observed number of administrations by the average time. We will determine carbon footprint by performing a life-cycle assessment for each magnesium formulation, then multiplying the observed number of administrations by the carbon footprint associated with that type of administration.

### Screening, enrollment, and participant timeline

Screening will be automated using the EHR (Appendix C). When an order for the electrolyte replacement protocol is placed for a patient who meets the inclusion criteria, an Epic OurPractice Advisory will appear on the screen stating “This patient meets inclusion criteria for the MAGNOLIA trial of higher-target versus lower-target protocolized magnesium replacement.” Below this statement, there will be the option to enroll or exclude (Figure 2). If exclude is chosen, the reason for exclusion must be specified as pre-eclampsia, sustained ventricular tachycardia, myasthenia gravis, or other. We will report the total number of ICU admissions during the study period, in order to quantify the proportion of patients for whom an electrolyte protocol was ordered.

**Figure 2:**
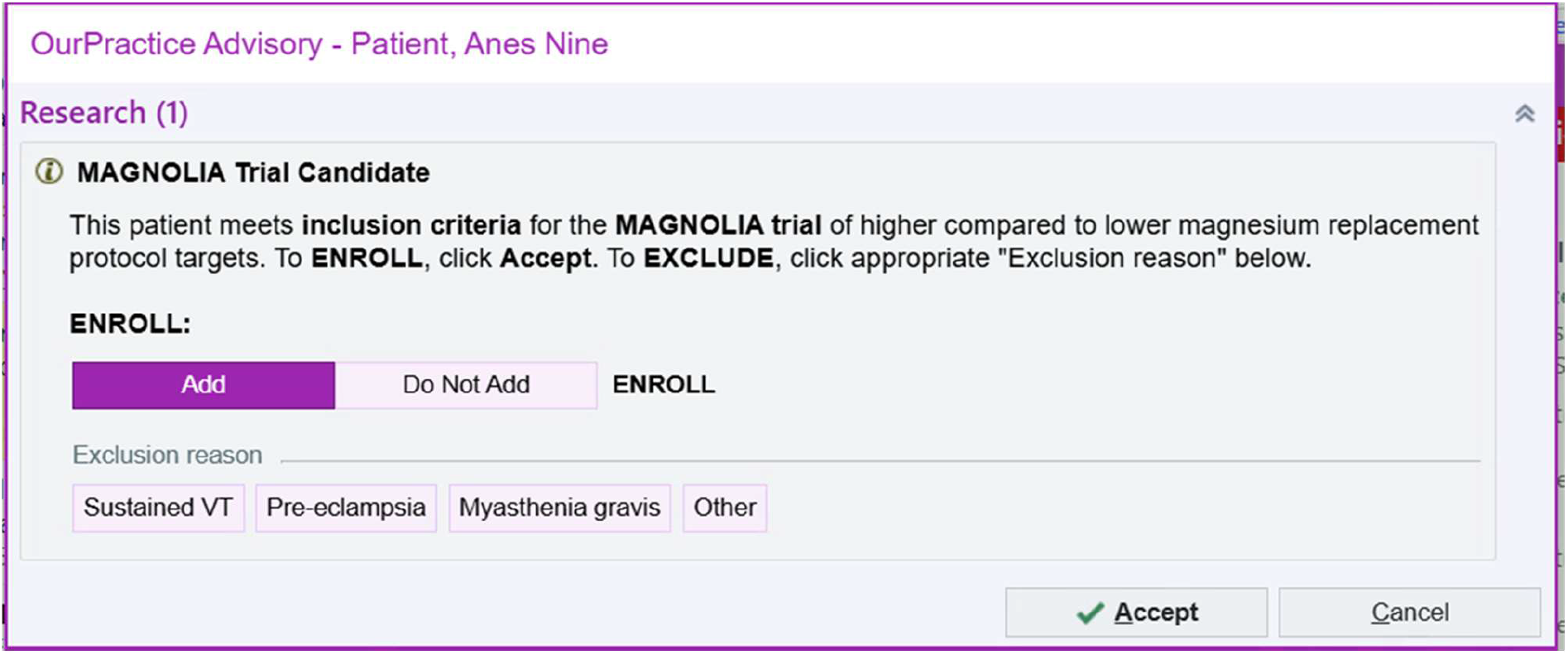
Electronic health record enrollment pop-up. Caption: This figure shows the pop-up alert that appears after clinicians sign orders for the critical care electrolyte replacement protocol on a patient who meets inclusion criteria. Clinicians are required to either enroll or exclude the patient in order to sign the electrolyte replacement protocol order.

Enrolled patients will be randomized and assigned as outlined in the “Assignment of interventions” section. For excluded patients, we will record the reason for exclusion, remove the patient from the trial, and the clinician will be free to order the usual institutional electrolyte protocol (higher target). Those who are withdrawn from the trial by patient or SDM withdrawal will be removed from analysis, while those who are withdrawn from the trial due to a medical decision by their treating physician will still be included in analyses according to an intention-to-treat paradigm. We will record the reasons why physicians withdraw patients from the trial.

### Sample size {19}

To calculate sample size, we performed simulations using an ordinal outcome with 5 levels (instead of the 32 levels in the primary outcome, because of small numbers of patients in many of the categories when using the 32-level outcome) and analyzed the simulations with proportional odds regression. We used frequentist proportional odds regression to speed up simulation, although the primary analysis of the trial data will use Bayesian regression. Based on data from participating sites, we assumed that 25% of participants would die, 5% of survivors would fall into each of the three middle levels corresponding to survival with different numbers of days with atrial fibrillation in ICU, and that 60% would survive to 30 days without having atrial fibrillation or flutter in the ICU.

To our knowledge, there is no established minimum clinically important difference (MCID) in days free of atrial fibrillation or flutter. In a systematic review of 78 trials enrolling critically ill patients that were published in high-impact journals, MCIDs for absolute reduction in mortality estimated by a panel of 10 experts ranged from 1% to 5%.(46) Using a five-point ordinal outcome with baseline probabilities as described above, an intervention with an odds ratio of 0.8 would result in outcome probabilities of 21%, 4.5%, 4.6%, 4.7%, and 65%. This would be a 4% absolute mortality reduction (from 25% to 21% of all patients) and a 2.5% absolute reduction in atrial fibrillation or flutter (from 15% to 12.5% of all patients).

Simulations showed that a trial of 3,253 patients would have 90% power to detect an odds ratio of 0.8, with a false positive rate of 5% or less. However, after the first interim analysis, we performed an additional power calculation focused on the clinically important subgroup of patients without evidence of atrial fibrillation at baseline. These patients comprised 75% of the first 1000 patients recruited. At 3,253 patients total, of whom 2,440 (∼75%) have no evidence of atrial fibrillation at baseline, there would be 73% power to detect an odds ratio of 0.8 with a false positive rate of 5% or less in this subgroup. At 3,853 patients, we would have an 80% power in this subgroup (and 93% power overall) to detect an odds ratio of 0.8 with false positive rate of 5% or less. We therefore opted to increase the sample size to 3,853, which amounts to approximately 6 additional weeks of recruitment.

### Recruitment {20}

This pragmatic comparative effectiveness trial is fully embedded in the EHR. This will facilitate rapid enrollment. Scarborough Health Network (SHN) admits approximately 2,000 patients per year to the non-cardiac ICUs across its three sites, while Lakeridge Health (LH) admits approximately 1,500 patients per year to its ICUs.

## Assignment of interventions: allocation

### Sequence generation {21a}

The sequence will not be pre-generated. We will use the computerized binary random number generator available in Epic Systems software.(28)

### Type of randomization {21b}

Simple binary randomization, 1:1 ratio between arms. The EHR used does not permit pre-generated sequences or randomization blocks.

### Concealment mechanism {22}

The randomization assignment is generated at enrollment and cannot be ascertained beforehand.

### Implementation {23}

Randomization is done by the random number generator in the Epic Systems software. Computer code programmed into the EHR will assign the intervention based on the randomization.

## Assignment of interventions: Blinding

### Who will be blinded {24a}

None of the patients or clinicians will be blinded in this pragmatic trial. The primary analysis and any interim analyses will be performed by a statistician not involved in data cleaning, using blinded group labels.

### Procedure for unblinding if needed {24b}

Assignment will be visible in the EHR for clinicians and will be shared with patients or SDMs upon their request.

## Data collection and management

### Plans for assessment and collection of outcomes {25a}

Outcome data will be gathered from the EHR using Structured Query Language (SQL) queries. The outcome of atrial fibrillation or flutter is charted by the ICU nurses based on their observations of continuous telemetry. In a validation assessment of 215 measurements, charted cardiac rhythm had a sensitivity of 94% and a specificity of 91% for atrial fibrillation or flutter, compared to the gold standard of continuous telemetry.(13)

### Plans to promote participant retention and complete follow-up {25b}

To promote retention, we will provide materials (poster, pamphlet, website) that communicate the purpose and importance of the study.

### Data management {26}

Data management will be integrated with the EHR. SQL queries will be used to generate a deidentified data table of baseline characteristics (Table 2) and a deidentified table of outcomes (Table 3), where each patient is identified only by an encrypted study ID. This deidentified table will be maintained as a comma-separated values file on a secure enterprise shared drive with password-protected access limited to the analysis team, or on a secure password-protected laptop owned by the lead statistician. Data will be maintained for 10 years before its deletion, to allow for its potential integration into meta-analyses.

### Confidentiality {33}

All identifying participant information will remain within the EHR.

### Plans for collection, laboratory evaluation and storage of biological specimens for genetic or molecular analysis in this trial/future use {32b}

There are no plans to collect biological specimens for genetic or molecular analysis.

## Statistical methods

### Statistical methods for primary and secondary outcomes {27a}

A full statistical analysis plan will be finalized before any final analyses of trial outcomes begins. All analyses will use Bayesian regression with weakly skeptical prior distributions (normal(0,1) in log-odds space).(47) The primary outcome will be analyzed according to assigned treatment (intention to treat) using a proportional odds model.(48) Ordinal secondary outcomes will be analyzed with the same approach. Binary secondary outcomes will be analyzed with logistic regression, lengths of stay will be analyzed with a negative binomial model, and magnesium levels will be analyzed with linear regression.

### Who will be included in the analyses {27b}

All randomized participants will be included in the analyses according to their assigned group, except for participants who are withdrawn due to patient or SDM request.

### Interim analyses {28b}

We will perform an interim analysis after 1000 and 2000 patients have 30-day outcomes available, to assess for harm in either group assignment. This is especially relevant because our trial uses opt-out consent.

If the probability of one treatment being inferior to the other with respect to mortality or the primary outcome is more than 0.99 in an interim analysis, then the trial will stop. If this occurs, immediately after the trial stops then all currently enrolled patients will be flipped to the protocol deemed superior.

If the probability of a difference in treatments is 0.90 or more but less than 0.99, the data monitoring committee will examine secondary analysis and outcome data to determine whether the trial should stop, suspend enrolment until the outcomes of all currently enrolled patients are known, or continue. If the data monitoring committee opts to recommend continuation, they may suggest an additional or earlier interim analysis than initially planned, to help guard against ongoing enrolment when one treatment has been demonstrated to be better than the other.

If the probability of a difference in treatments is less than 0.90, it is anticipated that the data monitoring committee will recommend continued enrollment.

### Methods for additional analyses (e.g. subgroup analyses) {27d}

We will perform pre-specified subgroup analyses modeling the interaction between subgroup membership and treatment effect with Bayesian regression. Pre-specified subgroups will include: sex (male vs female), age (quintiles), past history of atrial fibrillation or not, primary diagnosis of acute coronary syndrome or not, receiving vasopressors at time zero or not, invasively ventilated at time zero or not, end-stage renal disease at time zero or not, and primary diagnosis of sepsis or not. Credibility of subgroup effects will be assessed using the Instrument to assess the Credibility of Effect Modification ANalyses.(49) We also plan to perform risk-based and effect-based heterogeneity of treatment analyses and health economic analyses, with full details to follow in separate statistical analysis plans.(50)

### Methods in analysis to handle protocol non-adherence and any statistical methods to handle missing data {27c}

We will not impute missing outcome data. For heterogeneity of treatment effect modeling, we will impute missing baseline covariates using multiple imputation with chained equations.(51) The primary analysis will follow an “intention to treat” principle and will not adjust for protocol non-adherence.

### Plans to give access to the full protocol, participant level-data and statistical code {5}

We will post the protocol as a preprint and publish it in a peer-reviewed journal. All statistical code will be made available in a public repository with a digital object identifier (DOI). We will seek approval from the research ethics board to post a deidentified version of the data on the Health Data Nexus, a data platform allowing credentialed access to deidentified data supervised by the University of Toronto (https://healthdatanexus.ai/about/). (52)

## Oversight and monitoring

### Composition of the coordinating centre and trial steering committee {3d}

The day-to-day management of the trial will be performed by the SHN ICU research team. A trial steering committee, including the two principal investigators (CJY, TB), will meet monthly and as-needed to discuss planning, implementation, and conduct of the trial. All participating health networks will have representation on the steering committee.

### Composition of the data monitoring committee, its role and reporting structure {28a}

Because participants are enrolled without prior informed consent, we will use a data monitoring committee to provide an additional layer of independent oversight. The data monitoring committee will have a separate charter following published guidelines.(53) Its responsibility will be to ensure that patients enrolled in MAGNOLIA are not exposed to unnecessary or unreasonable risks, and that the trial is conducted according to the highest scientific and ethical standards.

It will be composed of two critical care medicine experts (national and international) and one statistician with Bayesian expertise. The data monitoring committee will meet and review data after each interim analysis, and as needed, in order to provide recommendations to the trial steering committee regarding continued enrolment and any other aspect of the trial pertinent to patient safety and research conduct.

### Adverse event reporting and harms {17}

Critically ill patients are admitted to the ICU for life-sustaining therapies, such as mechanical ventilation and vasopressors. Death and other serious complications are common in patients with critical illness. Due to these morbidity and mortality expectations in critically ill patients, and the intervention’s safety profile, only serious adverse events (SAEs) will be collected and reported The one exception will be hypermagnesemia: we will define a magnesium level of > 2.0 mmol/L as an adverse event and report its frequency.

We define a serious adverse event as any adverse medical occurrence that is believed by the investigators to be causally related to the intervention and results in any of the following: life-threatening condition (immediate risk of death); severe or permanent disability, prolonged hospitalization, or a significant hazard. SAEs will be reported to the data monitoring committee and the research ethics board.

To aid in SAE monitoring, we will use the established safety reporting software and database system at each participating hospital. These systems allow any team member to report potential safety incidents. We will query all safety incidents reported in these hospital safety reporting databases at participating sites during the time when the trial was active at that site. We will examine any that are assigned to Critical Care Departments or take place in a participating ICU and pertain to magnesium administration. For any such events, we will determine whether the affected patient was enrolled in MAGNOLIA (between day 0 and 30 of enrollment) at the time of the incident. If so, we will review the incident report and any relevant associated follow-up documentation to determine whether this represents a serious adverse event.

### Frequency and plans for auditing trial conduct {29}

We plan to audit magnesium levels and magnesium administrations in each arm. This audit will be performed every month for the first three months. If average magnesium levels differ by less than 0.1mmol/L between arms, or if protocol adherence is less than 70% in either arm, we will provide feedback to ensure clinicians understand the distinct protocols. In observational data from SHN, the observed protocol adherence in patients with magnesium levels near the current target of 0.95mmol/L was approximately 80%.

We will also track engagement with study materials: the number of visits to the study website, the number of times that QR codes on study materials are scanned, and whether a participant’s MyChart account is accessed after enrolment (because study information is available on MyChart).

### Plans for communicating important protocol amendments to relevant parties (e.g. trial participants, ethical committees) {31}

In the event of protocol modifications (eg, changes to eligibility criteria, outcomes, analyses), we will update trial registries, data monitoring committee, and investigators, and file an amendment to the research ethics board.

## Dissemination plans {8}

We plan to disseminate the trial results through publication and presentation at conferences, and seek to have the results publicized through the health network communications department. We will present the results at a meeting of the Patient-Family Advisor Committee or equivalent at each participating health network.

## Discussion

The MAGNOLIA trial is a comparative effectiveness RCT embedded into the EHR that seeks to compare magnesium replacement protocols targeting higher (>0.95mmol/L) and lower (>0.7mmol/L) serum levels in adult critically ill patients. This study will be embedded into the EHR, generating equitable, generalizable, secure, and pragmatic knowledge to address a key uncertainty in the care of critically ill patients.

Embedding the trial into the EHR strengthens equity and generalizability, increases efficiency, improves data security, and speeds post-study implementation of best practices. Equity and generalizability are strengthened because the automated screening process ensures that all patients for whom this is a clinically relevant question (ie patients for whom the magnesium replacement protocol is ordered) are able to enter into the study, regardless of sociodemographic or medical factors.(54,55) Efficiency is increased both by automating screening and by capturing outcomes from secondary data. Data security is improved because there is no requirement for sending information to an external database for randomization or to record study data. Post-study implementation of best practices will be straightforward, because we will maintain only the protocol which led to the better clinical outcomes (or the lower target, in the event of no difference).

The main drawback of EHR integration is that the number of sites who participate in the trial may be limited, restricting sample size. Not all sites have the ability to integrate trials into the EHR, because the feasibility of integration varies by site, by EHR, and according to local technical expertise. The MAGNOLIA trial addresses this limitation because it is built within one of Canada’s largest shared EHRs, which covers a multi-hospital catchment of 2500 inpatient beds.

The trial has other limitations. The participating sites do not care for many patients suffering critical illness related to trauma, cardiac surgery, neurosurgery, or transplant. The composite outcome is relevant to a critically ill population, but even if benefit is shown for the primary study outcome, subsequent studies will have to investigate whether a reduction in atrial fibrillation and flutter while hospitalized translates to improved longer term outcomes. The data itself will be limited by the accuracy of data entered for usual clinical care, which strengthens the pragmatism of the study but decreases the probability of identifying a difference in outcomes between the protocols.

In conclusion, the MAGNOLIA trial seeks to address an important question in critical illness, in a pragmatic fashion, using innovative EHR integration. It will enroll many patients from the Scarborough Health Network and Lakeridge Health, who have very rarely had the opportunity to participate in globally practice-changing critical care research. By seamlessly integrating research and quality improvement, our trial will set an example for aspiring learning healthcare systems worldwide.

## Trial status

Protocol version number: v1.3

Date: 2026-07-14

Recruitment commenced 25 November 2025

Recruitment completion estimated for June 1 2027

## Abbreviations

Atrial fibrillation: atrial fibrillation and flutter
EHR: electronic health record
ICU: intensive care unit
LH: Lakeridge Health
OR: odds ratio
MAGNOLIA: Effect of protocolized MAGNesium replacement in critically ill patients On mortaLIty and Atrial fibrillation: the MAGNOLIA randomized controlled trial
RCT: randomized controlled trial
SAE: serious adverse event
SDM: substitute decision maker
SHN: Scarborough Health Network

## Declarations

### Acknowledgements

Thanks to Dr Eddie Qian for his guidance regarding Epic coding, and Dr Blair Bigham for his advice on the posters.

### Authors’ contributions {31b}

CJY and TB are the chief investigators; they conceived the study and led the proposal and protocol development. CJY, KL, GT developed the statistical analysis plan. All authors contributed to study design, development of the proposal, and read and approved the protocol.

### Availability of data and materials {6}

We plan to make deidentified data available through Health Data Nexus after the publication of the trial and related secondary analyses.

### Ethics approval and consent to participate {30}

We have obtained Clinical Trials Ontario Research Ethics Board approval (CTO#5548), site-level approval at the Scarborough Health Network, and will procure site-level approval at Lakeridge Health prior to commencing study activities at that site. The rationale for our waiver of informed consent with the option to opt-out is outlined in the “Approach to consent” section.

### Competing interests {7b}

All authors and steering committee members declare no financial or other competing interests.

## Appendix F: SPIRIT 2025 randomized trial protocol extended checklist

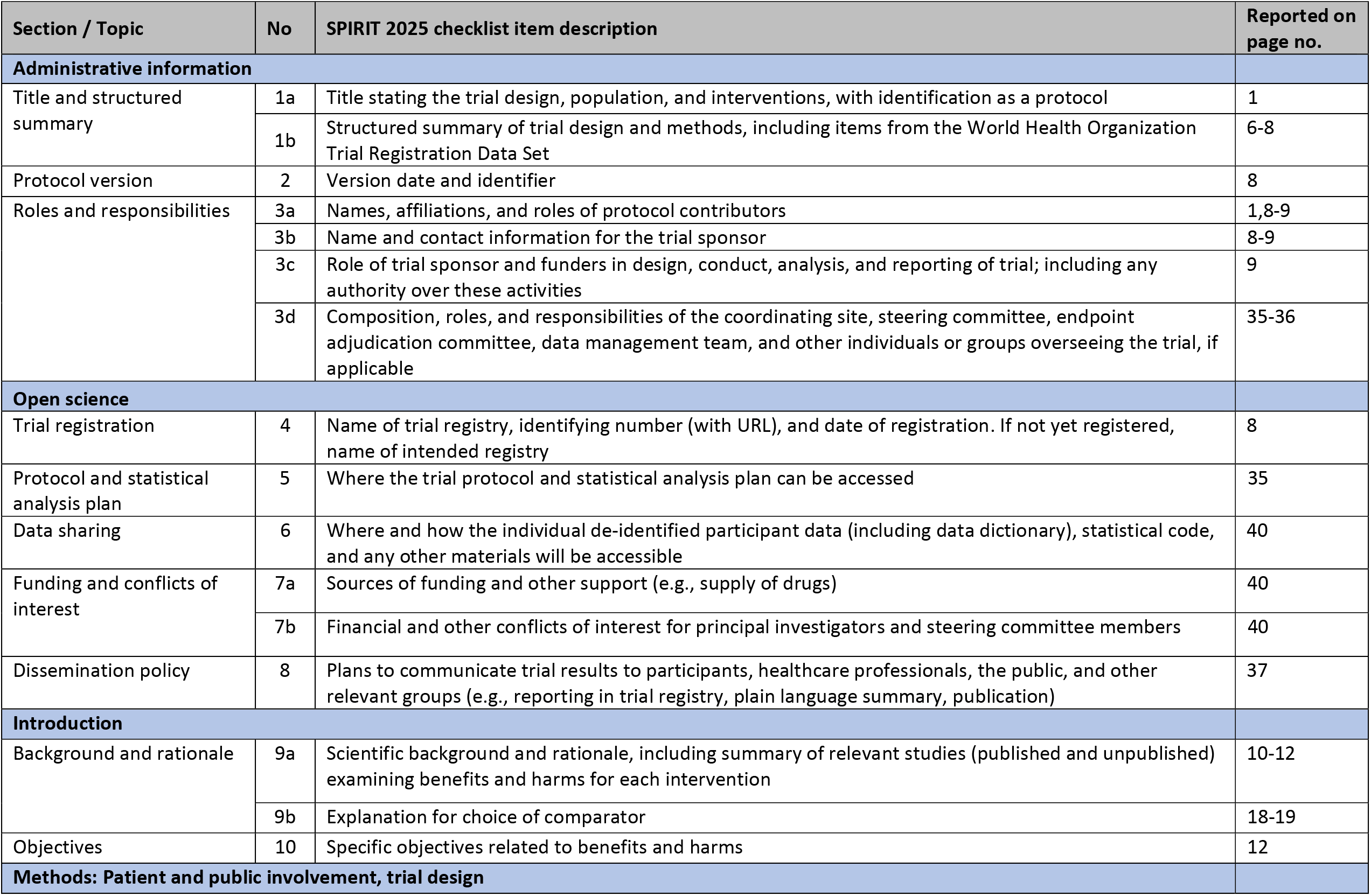
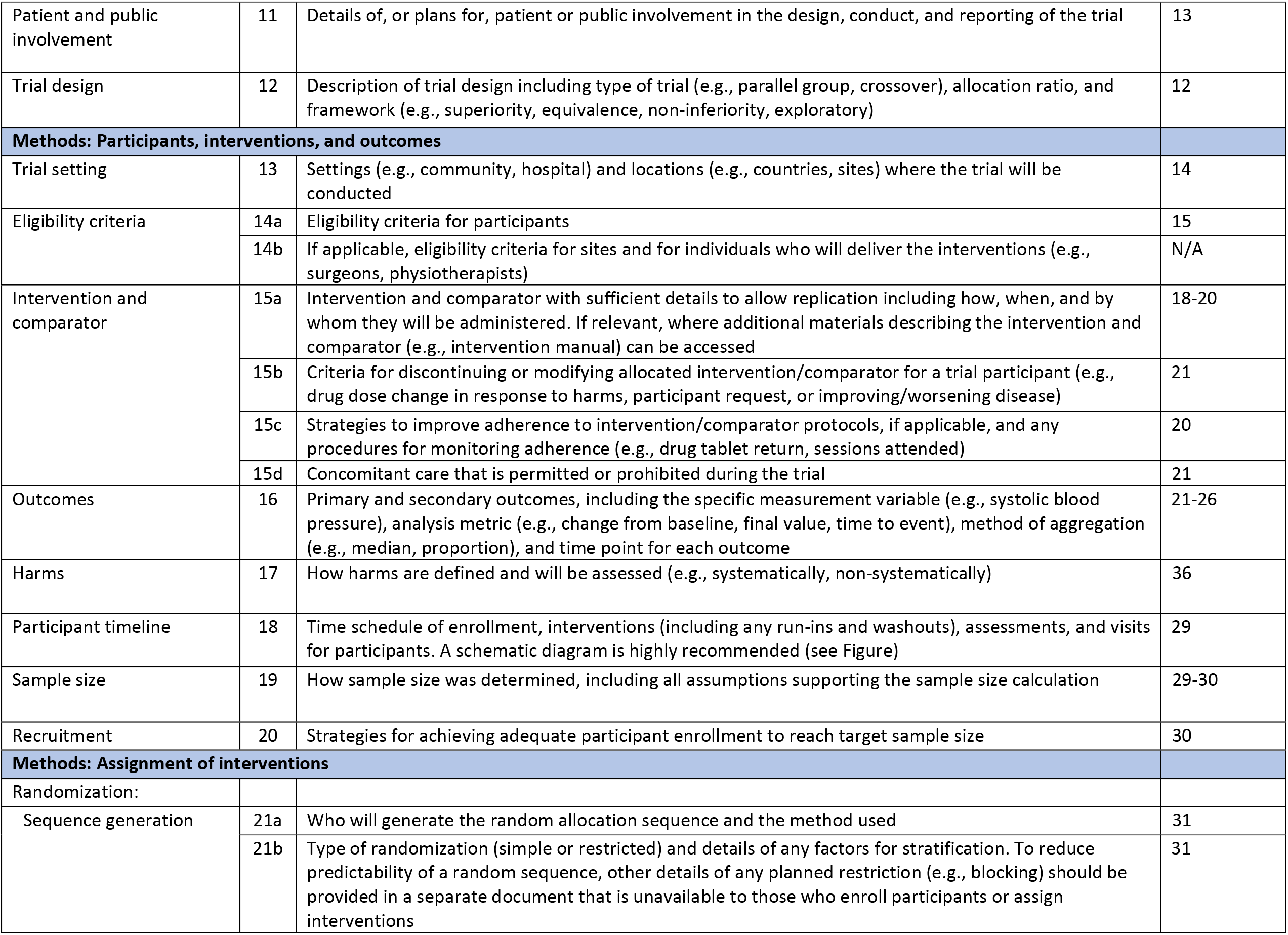
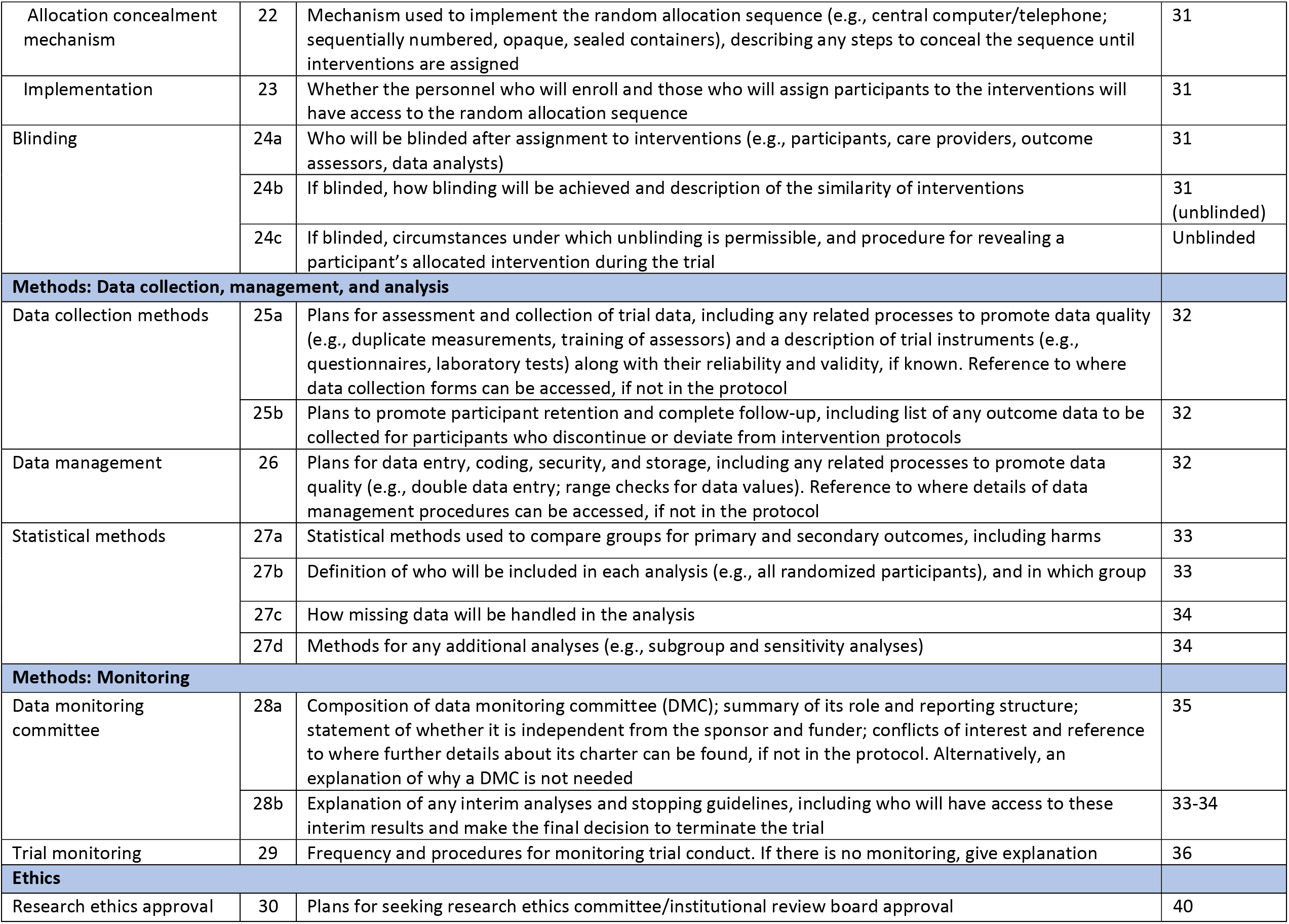
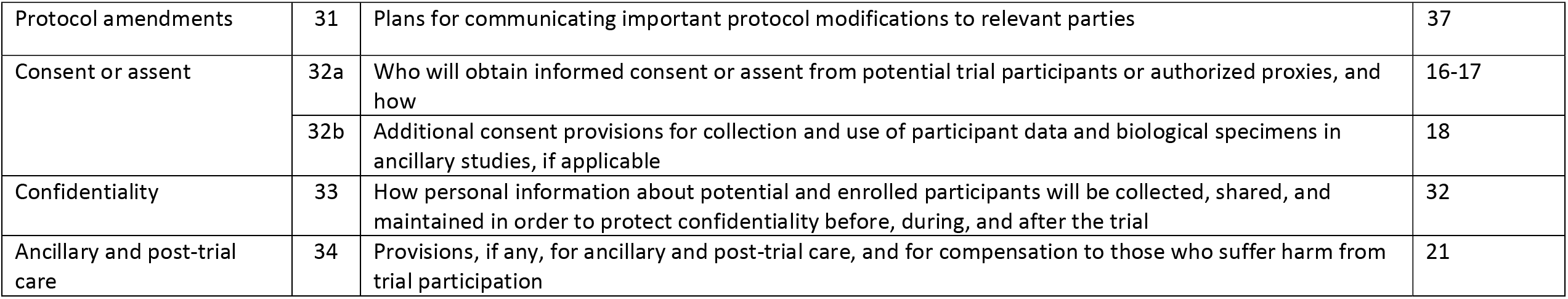

Citation: Chan A-W, Boutron I, Hopewell S, Moher D, Schulz KF, et al. SPIRIT 2025 statement: updated guideline for protocols of randomised trials. BMJ 2025;389:e081477. https://dx.doi.org/10.1136/bmj-2024-081477

© 2025 Chan A-W et al. This is an Open Access article distributed under the terms of the Creative Commons Attribution License (https://creativecommons.org/licenses/by/4.0/), which permits unrestricted use, distribution, and reproduction in any medium, provided the original work is properly cited.

## Supporting information

Protocol Appendix

Statistical Analysis Plan

## Data Availability

This clinical research protocol does not incorporate novel data.

