## Supplementary material for "Effect of protocolized MAGNesium replacement in critically ill patients On mortaLIty and Atrial fibrillation: protocol for the MAGNOLIA randomized controlled trial": Protocol Appendix

#### Names of protocol contributors

Christopher J Yarnell MD PhD
Joshua Craig, MSc
Selasi Bruce-Kemevor, RN
Lisa Burry PharmD PhD
Shannon Fernando, MD MSc
Rob Fowler, MDCM MSc
Keith Gunaratne, MD
Jan Haskings MEd
Francois Lamontagne, MD MSc
Kuan Liu PhD
Varuna Prakash MD
Stephanie Sibley MD
Karim Soliman, MD
George Tomlinson PhD
Louis Wei PharmD
Thomas Bodley MD MSc
For the Canadian Critical Care Trials Group

### Appendix A: Opt-out Procedure

#### Patient Contact / Outreach

The study team will make efforts to apprise patients of their enrollment in the trial (Figure 1). Having a continuous outreach strategy will ensure all patients have the opportunity to be aware of their enrollment and their right to opt-out.

At ICU admission: On their arrival to the ICU, patients will receive a pamphlet providing an overview of the SHN ICU (a pre-existing document unrelated to any studies). We will include an overview of the study as an insert in this pamphlet. The pamphlet will be provided to capable patients or to substitute decision makers (SDM) where patients do not have capacity. Bedside clinical staff will be educated on the study such that they can field basic questions and know to refer additional questions or requests to opt out to the research team. The study-specific pamphlet insert will include details of the study, a QR code to the study website, key contact information, and instructions on how to opt out.

During ICU admission: Posters will be displayed throughout the ICU and ICU waiting rooms containing an explanation of the trial, a QR code to the study website, key contact information, and instructions for opting out. Patients who are active on MyChart (or SDMs who use their loved ones' MyChart) will have the same information available in their account.

At any point during study conduct: Patients can visit the study website or contact the research team to review details about the study, explore answers to their questions, or opt out of the study. Patient portal (MyChart) will include information about the study indefinitely.

#### Opting Out

There are three ways a patient or SDM can reach the research team in order to opt-out:

During admission: At any point during admission, a patient or SDM can request to opt-out by speaking with a member of the research team or their clinical team. If this decision is communicated to the clinical team, the clinical team will let the research team know.

Via email: Patients or SDMs of incapable patients can email the research team to request that they opt-out. request to the research team via email alongside their name and date of birth. It is necessary to collect their name and date of birth to ensure the study team can correctly identify their chart and withdraw them from the trial. This email will be downloaded and stored in the study folder.

Via study website: The study website will contain research team contact information (email) with instructions to contact the research team should they seek to opt out on their own behalf or on the behalf of an incapable patient for whom they are a substitute decision maker.

When an opt-out request is received, the patient will be removed from the study. The patient / SDM will be posed the following open-ended questions, to explore whether their decision to opt-out is based on an easily-resolved misunderstanding:

1) Do you have any questions about the trial?
2) Do you have any questions about the opt-out model?
3) Is there a specific reason why you would like to opt-out?

Patients and their SDMs can decline to answer the above questions. After confirming the identity of the patient in question and their desire to opt out, we will verbally confirm to them that their data has been removed from the study. If the patient / SDM decides to remain in the study, perhaps as a result of clarification of misunderstandings via the questions above, they will remain enrolled.

If an opt-out request is received from a patient or substitute decision-maker, but we are unable to contact the patient or SDM to confirm the request, then we will remove the patient from the study.

### Appendix B: Patient Family Advisory Committee (PFAC) Consultation

**Date:** 19-June-2025

**Topic Presented:** “Calcium, Magnesium, Potassium, and Phosphate replacement inIntensive Care: The CaMPPhIre Research Program”

**Summary:** We presented the MAGNOLIA randomized controlled trial to the Scarborough Health Network’s (SHN) Patient and Family Advisory Council (PFAC). The PFAC is a volunteer group composed of individuals who have either been a patient at one of SHN’s hospitals, or are a family member of a patient.

#### Table 1: Attendance

| PFAC Committee and Members | SHN ICU Research Team |
|---|---|
| Gideon Quaison, Manager PFAC<br><br>Jan Hasking, Patient-family advisor<br><br>Patient and Family Partners: eleven (11) | Dr. Christopher Yarnell, MD PhD<br><br>Dr. Thomas Bodley, MD MSc<br><br>Joshua Craig, Research Coordinator<br><br>Selasi Bruce Kemevor, RN Res. Coordinator<br><br>Louis Wei, RPh, PharmD, SHN ICU<br><br>Elyse Eisner, Summer Research Student |

### Agenda

**Location:** Centenary Hospital - 2867 Ellesmere Road, Scarborough, ON M1E 4B9 (C3A-DR. BRUCE P. JOHNSTON CONFERENCE ROOM)
**Time:** 6:00 PM - 8:00 PM

**June 2025 PFAC Agenda**

| | Topic | Time | Presenters |
|---|---|---|---|
| | **Dinner** | 30 mins | N/A |
| 1 | **Welcome**<br><br>**Land Acknowledgement and African Ancestral Acknowledgement** | 2 mins | Gideon Quaison,<br>Manager, Health Equity, Patient & Community Engagement |
| 2 | **Connection Before Content** | 15 mins | Jan Haskings,<br>PFAC Co-chair |
| 3 | **Consultation & Feedback:** Calcium, Magnesium, Potassium, and Phospate replacement in Intensive Care: The CaMPPhIre Research Program | 25 mins | Dr. Christopher Yarnell<br>ICU and Critical Care Physician<br>Dr. Thomas Bodley<br>Physician |
| 4 | **(Second Study/Program)** | | |
| 5 | **Information Sharing** | 5 mins | Gideon Quaison |
| 6 | **Other Business** | 10 mins | All |
| 7 | **Wrap-Up** | 5 mins | Jan Haskings |

#### Specific Questions Posed to Groups (Three Breakout Groups):

> **SUMMARY**
>
> Background: Routine care is important for good outcomes in critical illness
>
> Goal: Find the best electrolyte protocol
>
> Method: Randomized trial built into Epic, with opt-out consent

1. What is your initial response to this?
2. What do you think of our proposed solution (opt out consent) to the problem of including all ICU patients?
3. What advice do you have on how to conduct this study in a way that preserves and builds trust with patients and families?

#### General Overview of Feedback

- Premise of study and research question were well received
- General questions about consent model
    - Patient and Family Partners (PFPs) understood the opt-out model and why it would be necessary
- PFPs highlighted the need for family education: about the study, the necessity of the opt-out model, and data security
    - PFPs suggested using active methods when possible (engaging the family at the bedside and distributing materials); passive information (posters, website), while necessary, may not be sufficient
    - PFPs noted that letting the patient / family know about the trial, their enrollment, and their choice to opt-out, sooner than later, is important
    - Including information about the study in ICU admission or discharge material could be a good failsafe measure to ensure all patients are captured

#### Group 1 Discussion and Feedback:

- Overall very well received by the members and they think it is something impactful for patients and could potentially lead to improved outcomes
- Opt-out consent:
  - There were many questions specifically regarding patients being automatically enrolled and only removed if opt'ed out
  - There was an understanding that due to clinical and logistical reasons, it might not be feasible to obtain consent at the time of ICU admission
  - Questions were raised about how the information will be provided to the patient/family members when the initial critical phase is over (and thus have the time/ability to consent)
    - Would the family/SDMs be informed their loved one was enrolled in the program and at that time? provided with the information and education for them to make a decision? Or, is the information simply passively available and thus family members/SDMs/patient might not even know about this or know of their freedom to opt out?
  - Questions were raised about how the patient/family would be educated, what kind of resource/education would be provided. The group felt it is important for this to happen.
- There were questions about how the patient's future care providers are informed (ward and GP). This was posed as a general question related to how the course of ICU care is communicated to these practitioners.
- Though we did not have a lot of time to discuss the 3rd question specifically, this theme was discussed in prior questions. Generally the consensus is "transparency"
  - The participants expressed they would be comforted knowing that the patient/family/SDM be provided the information (their loved one's enrolment in the trial, information on the trial and available options) sooner than later

#### Group 2 Discussion and Feedback:

- General sentiment of “What a common sense thing to do” among PFP participants
- There was a surprise that clinicians did not “already know the right levels” for electrolytes in the ICU
- PFPs mentioned “I like preventative as opposed to reactive” with respect to both therapy in the ICU as well as reaching out to patients/families regarding research
- The members wanted to understand if the data in the study would remain anonymous.
    - There was concern with opt-out consent if data was being accessed for research and that data would not be anonymized/deidentified.
    - Concern about cybersecurity
- Members highlighted the important to consider differences in trust in research / data across different cultures
    - Ensure multiple languages and reading level
    - It is helpful if people from community give out info
- People don’t have time / energy for a long consent process
- PFPs volunteered a suggestion that a poster with QR code link to more information about the study would be a useful item. They felt “people in ICU will read it”

#### Group 3 Discussion and Feedback:

What was your initial response:

- PFPs were surprised that there is a study for electrolyte replacement
    - They didn't know it was a problem
    - They were surprised at the variation in practice among hospitals
    - They were surprised there wasn't a consensus
- PFPs were curious as to how the question arose
    - Dr. Bodley explained that it began with a QI investigation into the amount of blood work being done, and an intent to reduce the amount of blood work being done. This endpoint seemed to resonate with the PFP
- Deferred vs. Opt Out consent
    - There were questions about whether any form of consent discussion would take place
    - Joshua Craig (research coordinator) clarified the difference between 'deferred consent' and the 'opt out' model
    - Dr. Bodley explained that since every patient would be affected by the trial (treatment wise), their main decision is whether they want their data included or excluded, and that there should be a focus on ensuring awareness so that patients and families know about the study and their right to opt out
        - PFPs suggested having some process for discussing the study with patients / families during the admission, when feasible
        - PFPs highlighted the importance of explaining data security to patients and families; that the data would be anonymized, etc.
        - PFPs highlighted that poster material might not be sufficient, not everyone will see a poster or interact with it
        - PFPs suggested including the study information and opt out instructions in the discharge package
- Connotation of 'trial'
    - PFPs discussed how 'trial' or 'study' connotes experimentation, which patients and families might be averse to participating in
    - Dr. Bodley explained that regardless of randomization, the intervention still operates within a standard of care
    - The PFPs seemed to appreciate this perspective but highlight the need to think about awareness materials, wording, etc.

Presentation Materials:

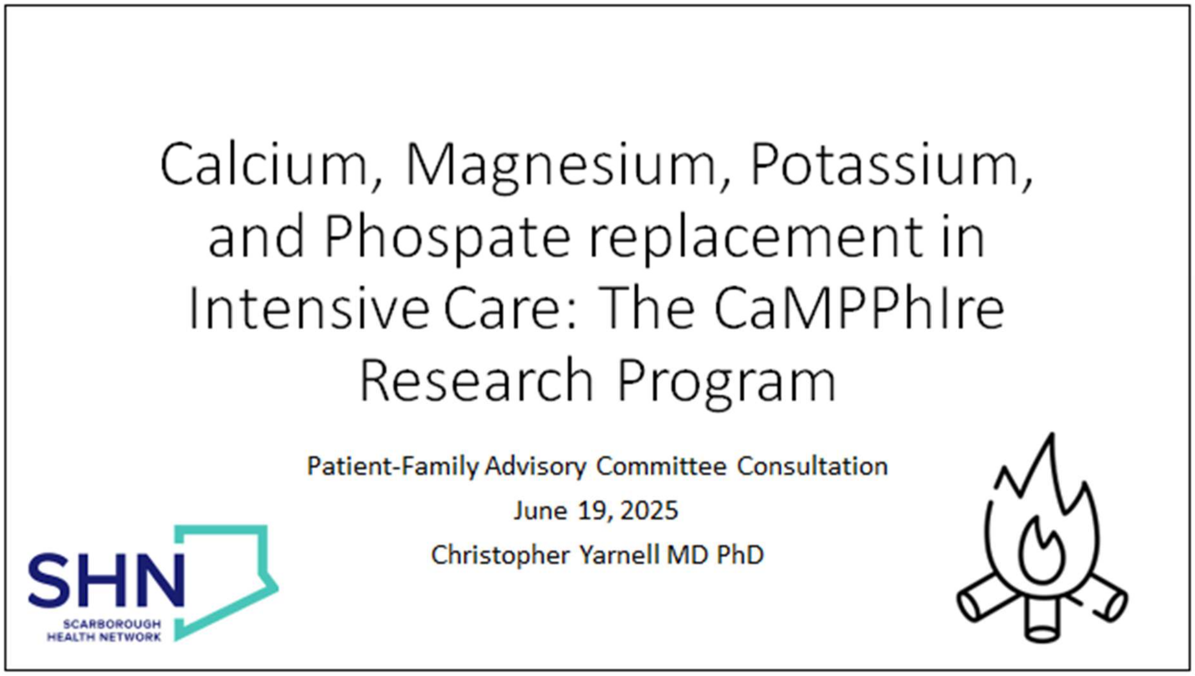

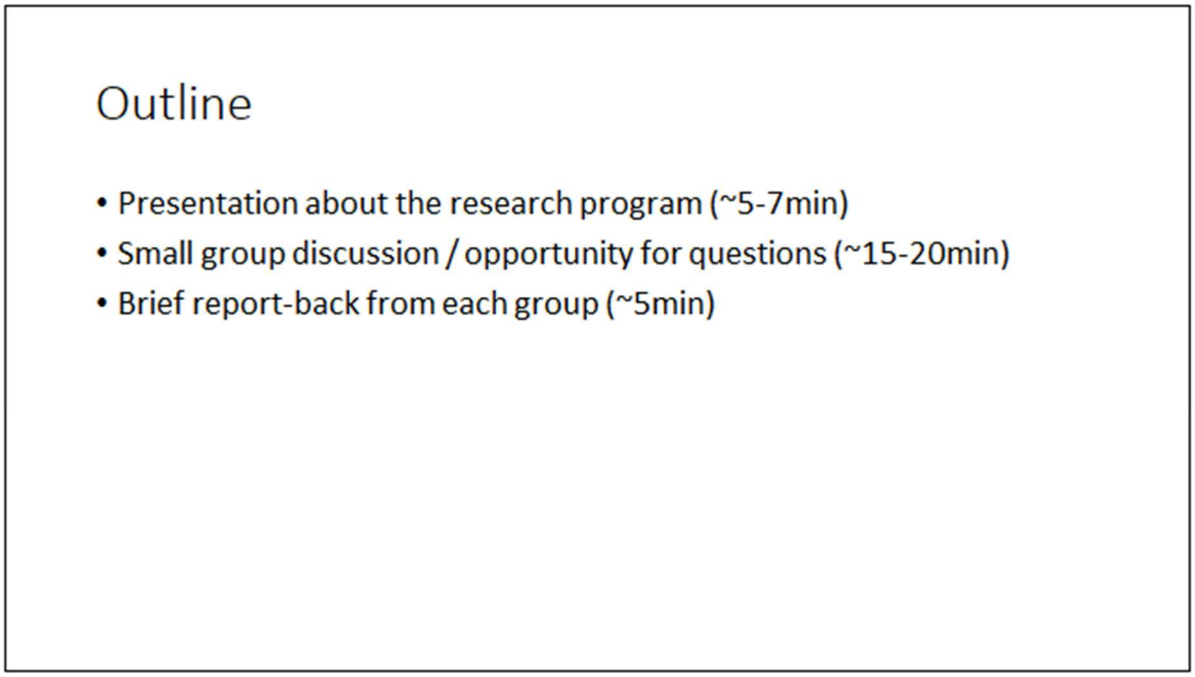

[Two slides deleted to remove photos of research team at MedrXiv request]

#### Problem: What to do about low electrolytes?

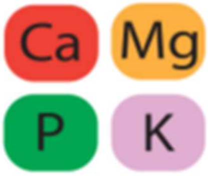

Calcium, magnesium, phosphate, potassium: often **low** in critically ill patients

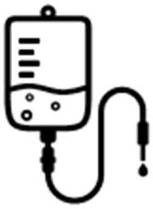

Protocols order intravenous doses when levels are low – **but protocols vary by institution**

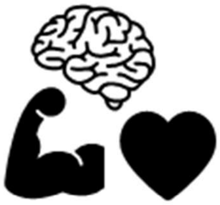

These electrolytes have important functions in brain, muscle, and heart

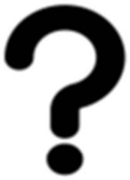

Which protocol is the best one, balancing benefits and harms? **It's unknown!**

#### Protocols are common and variable

**Normal range is 0.70-1.1 mmol/L**

| Hospital | Magnesium target |
|---|---|
| Scarborough Health Network | > 0.95 |
| Mount Sinai Hospital | > 0.90 |
| Amsterdam UMC (Netherlands) | > 0.90 |
| Toronto General Hospital | > 0.80 |
| Vancouver General Hospital | > 0.70 |
| Sunnybrook Hospital | > 0.60 |
| Nepean Hospital (Australia) | None |

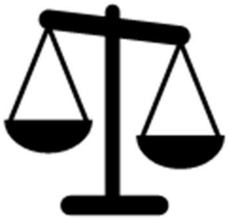

Which threshold best balances the **benefits** of maintaining electrolytes against the **harms and waste** of unnecessary doses?

#### The best way to learn this is with a randomized trial built into the electronic health record (EHR)

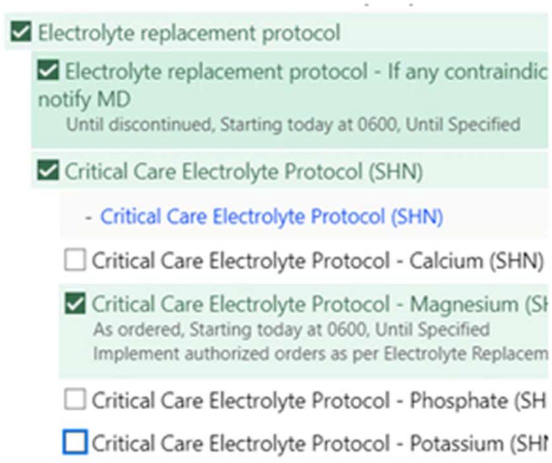

- Innovative
- Capitalize on our local advantages (biggest Epic implementation in Ontario)
- Efficient
- Equitable

#### Problem: 2000 critically ill patients per year

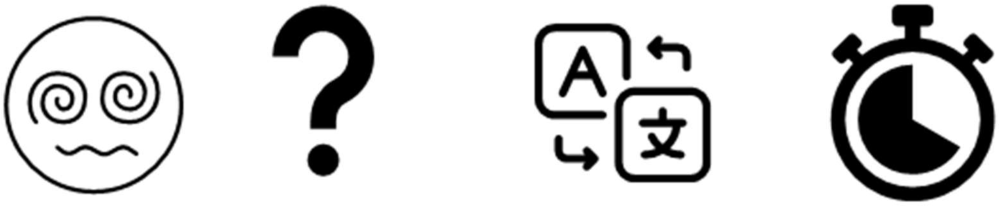

- Many are sedated, drowsy, confused
- Substitute decision makers can be unknown, unreachable, or **having the worst day of their lives**
- 30-40% prefer a non-English language (> 30 different languages)
- Some have very short stays (15-25% stay less than 2 days)

### Potential solution: Opt-out consent

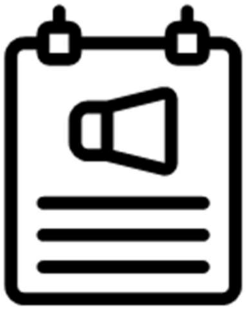

- Posters in ICU to let patients and families/caregivers know about study
- Option to opt out of data being included by emailing coordinator or letting bedside nurse know

### Questions for discussion

1. What is your initial response?
2. What do you think of our proposed solution (opt-out consent) to the problem of including all ICU patients?
3. What advice do you have on how to conduct this study in a way that **preserves and builds trust** with patients and families?

**SUMMARY**

Background: Routine care is important for good outcomes in critical illness

Goal: Find the best electrolyte protocol

Method: Randomized trial built into Epic, with opt-out consent

**THANK YOU!!**
**Please feel free to share thoughts and comments with, or with Jan or Gideon.**

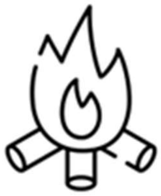

SHN
SCARBOROUGH HEALTH NETWORK

### Appendix C: Technical manual for embedding the trial into the electronic medical record

Christopher J Yarnell
Keith Gunaratne

#### Background

- Many aspects of routine care have no evidence to support their use
    - Significant variation exists across sites
    - In critical care medicine, many of the most important recent insights have shown that too much of an intervention for too long is harmful (tidal volume, SBTs, sedation)
- Routine care can be studied in observational or randomized fashion
    - Observational research rarely generates sufficiently convincing evidence to change practice
    - Randomized trials are also limited because they enroll non-representative populations, impose a high logistical and financial cost, and lack clear pathways for post-trial knowledge implementation
- Embedding an RCT into the EMR offers solutions to these problems
- Here we outline the technical implementation to accompany the MAGNOLIA trial protocol

#### Desired workflow

1. Screening, enrollment, randomization, assignment
    a. Physician orders magnesium replacement protocol on a patient as part of the ICU admission orders
    b. If patient is age 16 or older, a pop-up appears to notify the clinician of the patient's potential eligibility. It will have buttons for "Enroll" and for each of the three main exclusions
        i. If the clinician chooses an exclusion, the pop-up closes and the patient is noted to be ineligible in the system
        ii. If the clinician chooses to enroll, proceed
    c. The randomization and assignment happen instantly and the orders are signed, including the Critical Care Magnesium Electrolyte Protocol order
2. Ongoing care after MAGNOLIA enrolment or if patient is opted out
    a. The pre-existing workflow for the Critical Care Electrolyte Protocol remains the same (ICU RN clicks on the Critical Care Storyboard link after electrolyte results are available, and is guided through the electrolyte replacement orders by an order panel)

    b. Depending on the treatment assignment within the MAGNOLIA trial (lower target or higher target), the RN will be shown a different order panel corresponding to the assigned protocol
3. End of study intervention occurs at the earlier of:
    a. Critical Care Magnesium Electrolyte Protocol is discontinued (which is usual practice on ICU discharge)
    b. 30 full days elapse from enrollment (silent OurPractice Advisory will change patient MAGNOLIA study status to "Completed", and if the patient is still in ICU [or readmitted to ICU] with an active protocol after that, it will follow the pre-study baseline [higher target] protocol)

#### Study details

1. Study will be created as a Research Study using the Study Maintenance tool in Epic
    a. Title: Effect of protocolized MAGNesium replacement in critically ill patients On mortaLIty and Atrial fibrillation: the MAGNOLIA randomized controlled trial
    b. Principal investigator: Christopher Yarnell
    c. Additional investigators: Tom Bodley
    d. Research coordinators: Selasi Bruce-Kemevor, Joshua Craig
    e. Clinical trial registration: Clinicaltrials.gov
    f. REB approval: CTO5548

#### Interventions

- Each intervention will be an order panel built based on the current critical care electrolyte protocol order panel
- MAGNOLIA High-Target Protocol mirrors the current electrolyte protocol (replacement when magnesium level 0.95 mmol/L or less)
- MAGNOLIA Low-Target Protocol only mandates replacement when magnesium level is 0.7 mmol/L or less
- See trial protocol for specific details of the intervention magnesium replacement protocols

#### Trial screening, enrollment, randomization, and assignment

- An OurPractice Advisory will screen for eligible patients
    - Trigger: signing a critical care magnesium replacement protocol order
    - Criteria: Age 16 years or older, not already screened by the trial
    - Action: Present pop-up message: "This patient meets inclusion criteria for the MAGNOLIA trial of higher compared to lower magnesium replacement protocol targets."

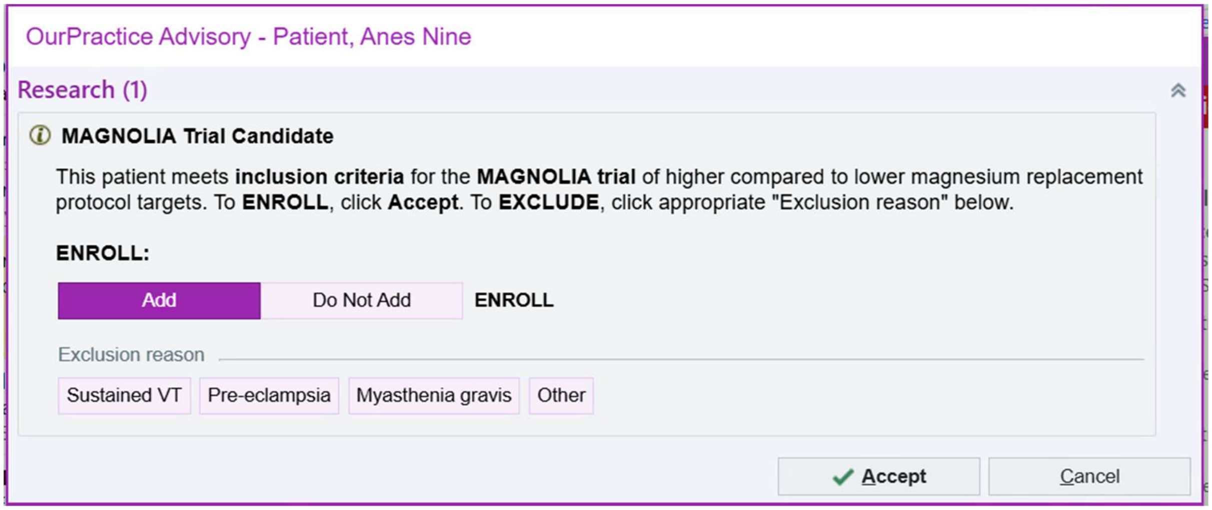

- OurPractice Advisory requires either enrollment (Add) or exclusion (Choose reason) before the advisory can be signed and orders can be processed
- If "Enroll" is selected, then:
    - Associate the patient to the study with status "Enrolled"
    - Randomize
    - Track enrolment and assignment in smartdata elements
- If "Exclude…" are selected
    - Associate the patient to the study with status "Ineligible"

#### Opt-out procedure

- If a patient opts out, the research team will change their study status to "Withdrawn"
- Patients will then be on the usual care (high target) protocol

#### Monitoring of protocol adherence

- SQL Caboodle report for number of magnesium levels according to assignment, whether supplementation recommended by protocol, and whether protocol was followed over 8 hours following the level.

#### Outcome ascertainment

- SQL Caboodle / Clarity query that pulls outcomes for each enrolled patient

### Edge cases

#### Patient enrolled and admitted to ICU but no electrolyte protocol ordered

- This is only possible if the electrolyte protocol is discontinued after ICU admission
- We will monitor for this in reports
- Based on clinical practice at participating sites, this is very rare, even in clinically stable patients

#### ICU readmission

- Electrolyte protocols are reordered on ICU readmission
- If an existing association with the MAGNOLIA study exists, and it is within 30 days from initial enrollment, then the bedside ICU nurse will be guided to the correct protocol

#### Transfer between participating hospitals

- Same as ICU readmission

#### John Doe / Duplicate MRN patients

- No robust way to prevent re-enrollment of previously enrolled patients if they come in and get registered under a different EPT (Patient) record.
- Once the merges of patient records are finished, this can be identified
- This is a rare issue
- In cases of multiple enrollments, the earlier enrollment will be the definitive one

#### Exclusion develops during study period

- If an exclusion develops during the study period such that the magnesium replacement protocol is contraindicated (eg confirmed diagnosis of myasthenia gravis), the protocol can be discontinued
- If a patient develops an indication for higher magnesium levels than those provided by the protocol (eg pre-eclampsia or ventricular tachycardia), the orders for additional magnesium can be placed in addition to the protocol

#### Enrolled/Randomized but never physically admitted to ICU

- These patients will be included

### Appendix D: Patient-facing materials and staff resource document

#### Pamphlet

**Scarborough Health Network**
**Critical Care Research**

SHN RESEARCH INSTITUTE

##### MAGNESIUM STUDY

###### What is Magnesium?

Magnesium is a mineral that helps your heart, muscles, & nerves function. Complications can occur when magnesium is too high or too low in the body.

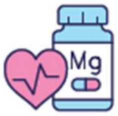

###### What is the study about?

- ICU patients are often given a magnesium supplement.
- Hospitals have different target magnesium levels for supplementation.
- We are conducting this study to learn which targets are best for patients to support the recovery of you or your loved one.

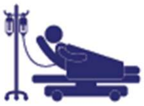

###### What is involved?

- All patients in the ICU are included.
- Patients are randomly assigned to a higher or lower magnesium target automatically.
- Both targets are within the standard of care (both would be considered safe and appropriate treatment options).
- We will find the best target by comparing cardiac health and patient outcomes between the two targets.

**Please Note:**
Study enrollment is automatic. If you do not want to be in the study or have questions, speak to your healthcare team or contact the ICU research team via email **** for assistance.

Scan QR code to learn more

மேலும் அறிய QR குறியீட்டை ஸ்கேன் செய்யவும்.
掃描二維碼以了解更多資訊

### Poster

[Poster deleted because it contained a picture of a member of the research team, at MedrXiv request]

Note that there are 6 versions of the poster above, which differ only in that they show 6 different members of the research team.

Staff resource (2 pages)

### MAGNESIUM STUDY (MAGNOLIA) | Staff Resource

SHN RESEARCH INSTITUTE

*This document was created to support ICU staff education & interactions with patients/ families in regards to this study*

#### Overview

- Protocolized magnesium replacement is common.
- The optimal target serum level for routine protocolized magnesium replacement is unknown.

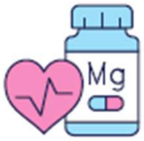

#### Objectives

- The goal of the study is to compare magnesium replacement strategies with different electrolyte protocols: one with replacement at Mg levels of 0.95, and the other at Mg levels of 0.70.
- Study team will evaluate burden of atrial fibrillation, hospital mortality, organ support (ventilator, vasopressor, and renal replacement therapy use), costs, nursing workload, and environmental impact.
- We want to find the magnesium target that best supports the recovery of patients.

#### How does it work?

- All patients admitted to the ICU are included unless the ICU Physician excludes the patient from the study.
- Patients are randomly assigned to a higher (≥ 0.95 mmol/L) or lower (≥ 0.70 mmol/L) magnesium replacement protocol via electronic medical record (EPIC).
- Protocols for both targets are within the standard of care **(there is no expected additional risk to patients by participating in the study)**.
- The protocol will instruct the bedside ICU nurse to administer Mg when serum Mg levels are below certain target (either 0.95 or 0.70).
- Patients on either protocol can receive additional magnesium ordered by a physician if the physician deems it necessary. However, the assigned protocol will remain unchanged.

#### What role is expected of bedside nursing / pharmacy?

- The research team needs your help. Patients or their family members may reach out to you with questions about the study (see FAQs next page).
- Patient study enrollment is automatic. However, **patients can opt out of the study at any time.**
- Opting out will change the magnesium protocol to our baseline pre-study protocol (with a higher magnesium threshold of 0.95). The patient's data will also be removed from the study.
- If a patient or patient SDM would like to be removed from the study, staff or patients/SDM may contact the ICU research team via email (****). Alternatively, staff may EPIC message research team members Joshua Craig and Nebadita Saha with the patient attached.

#### Questions?

### MAGNESIUM STUDY (MAGNOLIA) | Staff Resource

SHN RESEARCH INSTITUTE

#### Common Questions from Patients/Families

*This page includes common questions staff may receive from patients/families along with helpful responses*

##### Why is enrollment automatic?

The ICU is a fast paced high patient volume environment with complex patients and families that are often under extreme stress. It is not practical to collect direct consent from all patients / families in the ICU. The only practical way to ensure all patients have the opportunity to enroll in the study is to use an opt out approach.

##### How will I be informed about the study?

Both research & treatment teams will make every effort to ensure patient & families are informed about the study. Posters, pamphlets, and a website are available.

##### Is this study safe?

Both magnesium targets are within the standard of care, meaning there is no expected increased risk of participating. Individual patient care is always top priority, and the admitting physician or most responsible physician (MRP) can exclude patients from the study if they have concerns about enrollment. Additionally, patients will continue to receive all care that is indicated as per the physician regardless of study enrollment. We want to find the magnesium target that best supports the recovery of our ICU patients.

##### Who will have access to my information?

All study-related information will be kept safe and secure within the usual electronic medical record, (EPIC) accessible only to the clinical team and research representatives. Information that directly identifies you will **not** be disclosed or made publicly available.

##### How can I leave the study if I do not want to participate?

If a patient or patient SDM would like to be removed from the study, staff or patients/SDM may contact the ICU research team via email (****). Alternatively, staff may EPIC message research staff members Joshua Craig & Nebadita Saha with the patient attached.

##### Where can I learn more?

Patients/ families can scan the QR code on any magnesium study poster or pamphlet available on the unit to view the study website for more information. Additionally, they can email the ICU research team **** for specific questions or more information.

**Scan here for Mg Study website**

### Patient portal (MyChart) Information

The MyChart information displayed depends on the enrollment status of the patient. Patients will be either Enrolled (within 30 days from enrollment, not opted out), Withdrawn (opted out), Completed (more than 30 days from enrollment, opted out), or Ineligible (nothing displays in MyChart).

#### Study status = enrolled

**What is Magnesium?**

Magnesium (Mg) is a mineral that helps your heart, muscles, and nerves function. Complications can occur when magnesium is too high or too low in the body.

**What is the study about?**

- ICU patients are often given a magnesium supplement
- Hospitals have different target magnesium levels for supplementation
- We are conducting a study to learn which targets are best for patients

**Why does it matter?**

We want to find the magnesium target that best supports the recovery of you or your loved one.

**How the study works:**

1. All patients in the ICU are included, while they are in the ICU
2. Patients are randomly assigned to a higher or lower magnesium target automatically
3. Both targets are within the standard of care (the normal level of medical treatment)
4. We will find the best target by comparing cardiac health and patient outcomes between the two targets

*Please note: Study enrollment is automatic. If you do not want to be in the study or have questions, speak to the ICU team or contact the ICU research team via email for assistance.*

See https://www.shn.ca/magnesium-study/ for more information including Simplified Chinese and Tamil translations.

#### Study status = withdrawn

**You have opted out of the Magnesium Study. Please see below for additional information about the study, or if any questions.**

---

**What is Magnesium?**

Magnesium (Mg) is a mineral that helps your heart, muscles, and nerves function. Complications can occur when magnesium is too high or too low in the body.

**What is the study about?**

- ICU patients are often given a magnesium supplement
- Hospitals have different target magnesium levels for supplementation
- We are conducting a study to learn which targets are best for patients

**Why does it matter?**

We want to find the magnesium target that best supports the recovery of you or your loved one.

**How the study works:**

1. All patients in the ICU are included, while they are in the ICU
2. Patients are randomly assigned to a higher or lower magnesium target automatically
3. Both targets are within the standard of care (the normal level of medical treatment)
4. We will find the best target by comparing cardiac health and patient outcomes between the two targets

*Please note: Study enrollment is automatic. If you do not want to be in the study or have questions, speak to the ICU team or contact the ICU research team via email for assistance.*

See https://www.shn.ca/magnesium-study/ for more information including Simplified Chinese and Tamil translations.

#### Study status = Completed

**Thank you - you have completed the Magnesium Study. Please see below for additional information, or if questions.**

---

**What is Magnesium?**

Magnesium (Mg) is a mineral that helps your heart, muscles, and nerves function. Complications can occur when magnesium is too high or too low in the body.

**What was the study about?**

- ICU patients are often given a magnesium supplement
- Hospitals have different target magnesium levels for supplementation
- We are conducting a study to learn which targets are best for patients

**Why does it matter?**

We want to find the magnesium target that best supports the recovery of you or your loved one.

**How the study works:**

1. All patients in the ICU are included, while they are in the ICU
2. Patients are randomly assigned to a higher or lower magnesium target automatically
3. Both targets are within the standard of care (the normal level of medical treatment)
4. We will find the best target by comparing cardiac health and patient outcomes between the two targets

*Please note: Study enrollment was automatic. If would like your data removed from the study or have questions, speak to the ICU team or contact the ICU research team via email for assistance.*

See https://www.shn.ca/magnesium-study/ for more information including Simplified Chinese and Tamil translations.

### Appendix E: Research Ethics Board Approval

SHN RESEARCH INSTITUTE

**Date:** 1 October 2025

**To:** Dr. Christopher Yarnell, Scarborough Health Network

**Project ID:** ICU-25-018_5548

**Study Title:** Effect of protocolized MAGNesium replacement in critically ill patients On mortaLIty and Atrial fibrillation: the MAGNOLIA randomized controlled trial

**Study Sponsor:** Christopher Yarnell

**Application Type:** Clinical Trial Initial Application

**Review Type:** Full Board

**Meeting Date:** 01/Oct/2025

**Date Approval Issued:** 01/Oct/2025 12:04

**Study Approval Expiry Date:** 01/Oct/2026

---

Dear Dr. Yarnell,

Thank you for submitting the above referenced study on behalf of all participating sites through Stream. The Scarborough Health Network Research Ethics Board has reviewed the study and granted initial approval as of the date noted above.

**Study-wide documents approved:**

| Document Name | Document Date | Document Version |
|---|---|---|
| Data collection tools 2025-09-05 | 05/Sep/2025 | 1.0 |
| MAGNOLIA Pamphlet v1.0 5Sep2025 | 05/Sep/2025 | 1.0 |
| MAGNOLIA Poster SBK v1.0 5Sep2025 | 05/Sep/2025 | 1.0 |
| MAGNOLIA Staff Education v1.0 5Sep2025 | 05/Sep/2025 | 1.0 |
| MAGNOLIA WebsiteContent v1.0 5Sep2025.docx | 05/Sep/2025 | 1.0 |
| MAGNOLIA Staff Resource v1.0 2025-09-08 | 08/Sep/2025 | 1.0 |
| MAGNOLIA Study Pamphlet v1.0 2025-09-08 | 08/Sep/2025 | 1.0 |
| MAGNOLIA Study Poster v1.0 2025-09-08 | 08/Sep/2025 | 1.0 |
| MagnesiumReplacementRCT_Protocol v1.0 2025-09-05 | 05/Sep/2025 | 1.0 |
| MAGNOLIA Appendix A_ Opt out Procedure v1.0 5Sep2025 | 05/Sep/2025 | 1.0 |
| MAGNOLIA Appendix B_ PFAC Consultation v1.0 5Sep2025 | 05/Sep/2025 | 1.0 |
| MAGNOLIA Appendix C_ Technical Manual v1.0 5Sep2025 | 05/Sep/2025 | 1.0 |

**Study-wide documents acknowledged:**

| Document Name | Document Date | Document Version |
|---|---|---|
| MAGNOLIA Budget v1.0 5Sep2025.docx | 05/Sep/2025 | 1.0 |

Page 1 of 2

**Note: Study-wide REB approval does not confer ethics approval for participating sites. Each participating site, including that of the Lead Applicant, must submit the "Participating Site Initial Application" and receive approval from this REB prior to the conduct of the study at that site. All other required institutional approvals must also be obtained prior to the conduct of the study.**

No deviations from, or changes to, the protocol should be initiated without prior written approval from Scarborough Health Network Research Ethics Board, except when necessary to eliminate immediate hazard(s) to study participants or when the change(s) involves only administrative or logistical aspects of the trial (such as a change in telephone number).

REB members involved in the research project do not participate in the review, discussion or decision.

Scarborough Health Network Research Ethics Board operates in compliance with, and is constituted in accordance with, the requirements of the Tri-Council Policy Statement: Ethical Conduct for Research Involving Humans (TCPS 2); the International Conference on Harmonisation Good Clinical Practice Consolidated Guideline (ICH GCP); the WMA Declaration of Helsinki; Part C, Division 5 of the Food and Drug Regulations; Part 4 of the Natural Health Products Regulations; Part 3 of the Medical Devices Regulations and the provisions of the Personal Information Protection and Electronic Documents Act (PIPEDA) or the equivalent applicable provincial privacy law. Scarborough Health Network Research Ethics Board is qualified through the REB Qualification Program and is registered with the U.S. Department of Health and Human Services (DHHS) Office for Human Research Protection (OHRP).

Please do not hesitate to contact us if you have any questions.

Sincerely,

Sent on Behalf of Dr. Sherryn Roth

SHN REB Chair, MD

### Appendix F: SPIRIT 2025 randomized trial protocol extended checklist

| Section / Topic | No | SPIRIT 2025 checklist item description | Reported on page no. |
|---|---|---|---|
| **Administrative information** | | | |
| Title and structured summary | 1a | Title stating the trial design, population, and interventions, with identification as a protocol | 1 |
| | 1b | Structured summary of trial design and methods, including items from the World Health Organization Trial Registration Data Set | 6-8 |
| Protocol version | 2 | Version date and identifier | 8 |
| Roles and responsibilities | 3a | Names, affiliations, and roles of protocol contributors | 1,8-9 |
| | 3b | Name and contact information for the trial sponsor | 8-9 |
| | 3c | Role of trial sponsor and funders in design, conduct, analysis, and reporting of trial; including any authority over these activities | 9 |
| | 3d | Composition, roles, and responsibilities of the coordinating site, steering committee, endpoint adjudication committee, data management team, and other individuals or groups overseeing the trial, if applicable | 35-36 |
| **Open science** | | | |
| Trial registration | 4 | Name of trial registry, identifying number (with URL), and date of registration. If not yet registered, name of intended registry | 8 |
| Protocol and statistical analysis plan | 5 | Where the trial protocol and statistical analysis plan can be accessed | 35 |
| Data sharing | 6 | Where and how the individual de-identified participant data (including data dictionary), statistical code, and any other materials will be accessible | 40 |
| Funding and conflicts of interest | 7a | Sources of funding and other support (e.g., supply of drugs) | 40 |
| | 7b | Financial and other conflicts of interest for principal investigators and steering committee members | 40 |
| Dissemination policy | 8 | Plans to communicate trial results to participants, healthcare professionals, the public, and other relevant groups (e.g., reporting in trial registry, plain language summary, publication) | 37 |
| **Introduction** | | | |
| Background and rationale | 9a | Scientific background and rationale, including summary of relevant studies (published and unpublished) examining benefits and harms for each intervention | 10-12 |
| | 9b | Explanation for choice of comparator | 18-19 |

| | | | |
|---|---|---|---|
| Objectives | 10 | Specific objectives related to benefits and harms | 12 |
| **Methods: Patient and public involvement, trial design** | | | |
| Patient and public involvement | 11 | Details of, or plans for, patient or public involvement in the design, conduct, and reporting of the trial | 13 |
| Trial design | 12 | Description of trial design including type of trial (e.g., parallel group, crossover), allocation ratio, and framework (e.g., superiority, equivalence, non-inferiority, exploratory) | 12 |
| **Methods: Participants, interventions, and outcomes** | | | |
| Trial setting | 13 | Settings (e.g., community, hospital) and locations (e.g., countries, sites) where the trial will be conducted | 14 |
| Eligibility criteria | 14a | Eligibility criteria for participants | 15 |
| | 14b | If applicable, eligibility criteria for sites and for individuals who will deliver the interventions (e.g., surgeons, physiotherapists) | N/A |
| Intervention and comparator | 15a | Intervention and comparator with sufficient details to allow replication including how, when, and by whom they will be administered. If relevant, where additional materials describing the intervention and comparator (e.g., intervention manual) can be accessed | 18-20 |
| | 15b | Criteria for discontinuing or modifying allocated intervention/comparator for a trial participant (e.g., drug dose change in response to harms, participant request, or improving/worsening disease) | 21 |
| | 15c | Strategies to improve adherence to intervention/comparator protocols, if applicable, and any procedures for monitoring adherence (e.g., drug tablet return, sessions attended) | 20 |
| | 15d | Concomitant care that is permitted or prohibited during the trial | 21 |
| Outcomes | 16 | Primary and secondary outcomes, including the specific measurement variable (e.g., systolic blood pressure), analysis metric (e.g., change from baseline, final value, time to event), method of aggregation (e.g., median, proportion), and time point for each outcome | 21-26 |
| Harms | 17 | How harms are defined and will be assessed (e.g., systematically, non-systematically) | 36 |
| Participant timeline | 18 | Time schedule of enrollment, interventions (including any run-ins and washouts), assessments, and visits for participants. A schematic diagram is highly recommended (see Figure) | 29 |
| Sample size | 19 | How sample size was determined, including all assumptions supporting the sample size calculation | 29-30 |
| Recruitment | 20 | Strategies for achieving adequate participant enrollment to reach target sample size | 30 |
| **Methods: Assignment of interventions** | | | |
| Randomization: | | | |
| Sequence generation | 21a | Who will generate the random allocation sequence and the method used | 31 |

| | | | |
|---|---|---|---|
| | 21b | Type of randomization (simple or restricted) and details of any factors for stratification. To reduce predictability of a random sequence, other details of any planned restriction (e.g., blocking) should be provided in a separate document that is unavailable to those who enroll participants or assign interventions | 31 |
| Allocation concealment mechanism | 22 | Mechanism used to implement the random allocation sequence (e.g., central computer/telephone; sequentially numbered, opaque, sealed containers), describing any steps to conceal the sequence until interventions are assigned | 31 |
| Implementation | 23 | Whether the personnel who will enroll and those who will assign participants to the interventions will have access to the random allocation sequence | 31 |
| Blinding | 24a | Who will be blinded after assignment to interventions (e.g., participants, care providers, outcome assessors, data analysts) | 31 |
| | 24b | If blinded, how blinding will be achieved and description of the similarity of interventions | 31 (unblinded ) |
| | 24c | If blinded, circumstances under which unblinding is permissible, and procedure for revealing a participant's allocated intervention during the trial | Unblinded |
| **Methods: Data collection, management, and analysis** | | | |
| Data collection methods | 25a | Plans for assessment and collection of trial data, including any related processes to promote data quality (e.g., duplicate measurements, training of assessors) and a description of trial instruments (e.g., questionnaires, laboratory tests) along with their reliability and validity, if known. Reference to where data collection forms can be accessed, if not in the protocol | 32 |
| | 25b | Plans to promote participant retention and complete follow-up, including list of any outcome data to be collected for participants who discontinue or deviate from intervention protocols | 32 |
| Data management | 26 | Plans for data entry, coding, security, and storage, including any related processes to promote data quality (e.g., double data entry; range checks for data values). Reference to where details of data management procedures can be accessed, if not in the protocol | 32 |
| Statistical methods | 27a | Statistical methods used to compare groups for primary and secondary outcomes, including harms | 33 |
| | 27b | Definition of who will be included in each analysis (e.g., all randomized participants), and in which group | 33 |
| | 27c | How missing data will be handled in the analysis | 34 |
| | 27d | Methods for any additional analyses (e.g., subgroup and sensitivity analyses) | 34 |
| **Methods: Monitoring** | | | |
| Data monitoring committee | 28a | Composition of data monitoring committee (DMC); summary of its role and reporting structure; statement of whether it is independent from the sponsor and funder; conflicts of interest and reference | 35 |

| | | | |
|---|---|---|---|
| | | to where further details about its charter can be found, if not in the protocol. Alternatively, an explanation of why a DMC is not needed | |
| | 28b | Explanation of any interim analyses and stopping guidelines, including who will have access to these interim results and make the final decision to terminate the trial | 33-34 |
| Trial monitoring | 29 | Frequency and procedures for monitoring trial conduct. If there is no monitoring, give explanation | 36 |
| **Ethics** | | | |
| Research ethics approval | 30 | Plans for seeking research ethics committee/institutional review board approval | 40 |
| Protocol amendments | 31 | Plans for communicating important protocol modifications to relevant parties | 37 |
| Consent or assent | 32a | Who will obtain informed consent or assent from potential trial participants or authorized proxies, and how | 16-17 |
| | 32b | Additional consent provisions for collection and use of participant data and biological specimens in ancillary studies, if applicable | 18 |
| Confidentiality | 33 | How personal information about potential and enrolled participants will be collected, shared, and maintained in order to protect confidentiality before, during, and after the trial | 32 |
| Ancillary and post-trial care | 34 | Provisions, if any, for ancillary and post-trial care, and for compensation to those who suffer harm from trial participation | 21 |

Citation: Chan A-W, Boutron I, Hopewell S, Moher D, Schulz KF, et al. SPIRIT 2025 statement: updated guideline for protocols of randomised trials. BMJ 2025;389:e081477. https://dx.doi.org/10.1136/bmj-2024-081477

© 2025 Chan A-W et al. This is an Open Access article distributed under the terms of the Creative Commons Attribution License (https://creativecommons.org/licenses/by/4.0/), which permits unrestricted use, distribution, and reproduction in any medium, provided the original work is properly cited.
