## Supplementary material for "Effect of protocolized MAGNesium replacement in critically ill patients On mortaLIty and Atrial fibrillation: protocol for the MAGNOLIA randomized controlled trial": Statistical Analysis Plan

Trial Registration: NCT07173855

[Statistical Analysis Plan Version 1.0 — 2026-10-08]

[Protocol Version 1.3 referenced — 2026-07-13]

### Names of protocol contributors

Christopher J Yarnell MD PhD (Co-principal investigator)
Zhengyang Fei MSc (Analyst)
Karen E. A. Burns MD MSc
Rob Fowler MDCM MSc
Kuan Liu PhD (Statistician)
Varuna Prakash MD
Edward Qian MD MSc
Karim Soliman MD
George Tomlinson PhD (Statistician)
Thomas Bodley MD MSc (Co-principal investigator)

### Section 1: Administrative Information

#### Item 1a. Title

Statistical Analysis Plan for the Effect of protocolized MAGNesium replacement in critically ill patients On mortaLIty and Atrial fibrillation (MAGNOLIA) randomized controlled trial

#### Item 1b. Trial registration number

NCT07173855

#### Item 2. SAP version number with dates

Version 1.0 — 2026-09-09

#### Item 3. Reference to version of protocol being used

Protocol Version 1.3 — 2026-07-13

#### Item 4a. SAP revision history

*List each revision with version number, date, and author.*

1. Version 1.0, 2026-09-09, Christopher Yarnell

#### Item 4b. Justification for each SAP revision

1. Initial protocol draft.

#### Item 4c. Timing of SAP revisions in relation to interim analyses, etc.

Initial draft completed after first interim analysis, during which principal investigators and steering committee members were blinded to results according to treatment assignment but able to review pooled demographic, adherence, and outcome data.

#### Item 5. Names, affiliations, and roles of SAP contributors

*Include Principal Investigator, senior/responsible statistician, and other contributors with affiliations.*

Christopher J Yarnell, MD PhD (Co-principal Investigator) - Department of Critical Care Medicine and Research Institute, Scarborough Health Network, Toronto, Canada; Department of Medicine and Interdepartmental Division of Critical Care Medicine, University of Toronto, Toronto, Canada

Zhengyang Fei, MSc – Scarborough Health Network Research Institute, Toronto, Canada

Karen E. A. Burns, MD MSc – Department of Critical Care Medicine, Unity Health Toronto; and Interdepartmental Division of Critical Care Medicine, University of Toronto, Toronto, Canada

Rob Fowler, MDCM MSc – Sunnybrook Health Sciences Centre, Toronto, Canada; Department of Medicine and Interdepartmental Division of Critical Care Medicine, University of Toronto, Toronto, Canada

Kuan Liu PhD (Statistician) – Institute of Health Policy, Management, and Evaluation, University of Toronto, Toronto, Canada

Varuna Prakash MD – Lakeridge Health Department of Critical Care Medicine, Oshawa, Canada; Department of Critical Care Medicine, Queen's University, Kingston, Canada

Karim Soliman, MD – Lakeridge Health Department of Critical Care Medicine, Oshawa, Canada; Department of Critical Care Medicine, Queen's University, Kingston, Canada

Edward Qian, MD MSc – Division of Allergy, Pulmonary, and Critical Care Medicine, Vanderbilt University Medical Center, Nashville, United States of America; Department of Biomedical Informatics, Vanderbilt University Medical Center, Nashville, United States of America

George Tomlinson PhD (Statistician) – University Health Network, Toronto, Canada; University of Toronto, Toronto, Canada

Thomas Bodley, MD MSc (Co-principal Investigator) – Department of Critical Care Medicine and Research Institute, Scarborough Health Network, Toronto, Canada; Department of Medicine and Interdepartmental Division of Critical Care Medicine, University of Toronto, Toronto, Canada

**Item 6a. Signature: person writing the SAP**

Christopher Yarnell

**Item 6b. Signature: senior statistician responsible**

George Tomlinson

**Item 6c. Signature: chief investigator / clinical lead**

Christopher Yarnell, MD PhD

### Section 2: Introduction

#### Item 7. Synopsis of trial background and rationale

Magnesium is essential for cellular function, with important roles in regulating cardiac rhythm, vascular tone, and the nervous system.[1] Low magnesium levels are common in critically ill patients, and clinical consequences include cardiac dysrhythmias such as atrial fibrillation and flutter.[2] Meta-analyses of randomized trials show that intravenous magnesium may prevent atrial fibrillation after cardiac surgery[3–5], however, in non-cardiac critically ill patients, the effects are less certain.[6,7] No studies have evaluated the common practice of protocolized magnesium administration based on serum levels (replacement) in critically ill patients.[8]

Higher target magnesium levels (e.g. 0.95 mmol/L) increase the likelihood of replacement and may improve mortality or reduce atrial fibrillation.[6,7] Observational studies conflict on whether magnesium supplementation reduces tachyarrhythmia.[9,10] Lower target levels (e.g. 0.70mmol/L) lead to fewer doses of electrolytes, which reduces related fluid administration[11,12], costs[12], nursing workload[13], and environmental waste.[11] Avoiding unnecessary fluid administration is important, because positive cumulative fluid balance is associated with mortality.[14]

Given the implications for patients, the environment, and costs, it is crucial to investigate whether protocolized magnesium replacement aiming at a higher (0.95 mmol/L) or lower (0.7 mmol/L) target leads to lower incidence of mortality and atrial fibrillation in critically ill patients. To this end, we proposed the MAGNOLIA RCT, and in this document describe the Statistical Analysis Protocol (SAP) according to a recommended SAP checklist.[15]

#### Item 8. Description of specific objectives

To compare protocolized magnesium replacement targeting higher (magnesium > 0.95 mmol/L) and lower (magnesium > 0.7 mmol/L) levels in adult patients with critical illness, with respect to:

1) Burden of atrial fibrillation
2) Hospital mortality
3) Organ support (ventilator, vasopressor, and renal replacement therapy use)

### Section 3: Study Methods

#### Item 9. Trial design overview

Pragmatic, superiority, parallel-group, randomized controlled trial embedded in the electronic health record, comparing higher versus lower magnesium supplementation targets. Allocation ratio: 1:1. The primary outcome is an ordinal composite of mortality and days alive and free of atrial fibrillation and flutter in critical illness.

#### Item 10. Randomization details

Simple binary randomization without stratification, due to the constraints of the software allowing embedding of the trial into the electronic health record.

#### Item 11. Full sample size calculation

Sample size calculation is found on pages 30-31 of the protocol. We used simulation and a 5-category ordinal outcome with observed frequencies similar to the trial data. This showed that a trial of 3,853 patients had 93% power to detect an odds ratio of 0.8, with a false positive rate of 5% or less. Power was 80% in the important subgroup of patients without evidence of atrial fibrillation or flutter at baseline. We used the 5-category ordinal outcome because many ordinal levels have few patients in them, and we planned to analyze a 5-category ordinal outcome as a back-up if analyzing the full 32-level ordinal outcome failed. We assumed that power would be equal or greater when analyzing the full 32-level ordinal outcome.

#### Item 12. Hypothesis-testing framework and comparisons

The trial has adopted a Bayesian superiority framework, planning to report the probability that the higher target is superior to the lower target with 95% credible intervals (CrI).

#### Item 13a. Interim analyses

As described on page 33 of the protocol, we plan two interim analyses, after 1000 and 2000 patients have primary outcome information available. Adherence data, pooled demographic data, and pooled outcome data will be reviewed by the steering committee and the data monitoring committee. Demographic and outcome data by assignment, with assignment labels blinded, will be reviewed by the data monitoring committee, with the option for unblinding if needed to determine the best recommendation for the investigators and steering committee.

#### Item 13b. Any planned adjustment of the significance level due to interim analysis

In this Bayesian trial, no significance level adjustments are planned.

#### Item 13c. Details of guidelines for stopping the trial early

The data monitoring committee has been tasked to recommend stopping if the probability of superiority with respect to the primary outcome or mortality is > 0.99 or < 0.01. If the probability of superiority with respect to the same outcomes lies between 0.90 and 0.99, or between 0.01 and 0.10, then they will incorporate secondary outcome information into their decision whether to recommend continuation, stopping, or additional interim analyses.

#### Item 14. Timing of final analysis

Final primary analysis will be completed after the primary outcome is available for all enrolled patients, and final analysis of secondary outcomes ascertained at 30, 60, and 90 days will be completed after they are available.

#### Item 15. Time points at which outcomes are measured

Outcomes will be assessed at 30, 60, and 90 days after enrollment.

### Section 4: Statistical Principles

#### Item 16. Level of statistical significance

We will report the probabilities of superiority and the point estimates of odds ratios with 95% credible intervals for primary and secondary analyses.

#### Item 17. Description and rationale for any adjustment for multiplicity

No adjustments for multiplicity are planned. All secondary outcomes will be considered exploratory, and whether their findings align with the primary analysis will be considered as part of the overall interpretation of the trial results.

#### Item 18. Confidence intervals to be reported

We will quantify uncertainty with 95% credible intervals (CrI).

#### Item 19a. Definition of adherence to the intervention and how this is assessed, including extent of exposure

We will define adherence as whether the recommended action (magnesium supplementation administered or not), according to the assigned protocol, is taken within 8 hours following a serum magnesium level becoming available, in an enrolled patient with an active magnesium protocol ordered who is within 30 days of enrollment. We will also report the proportion of intensive care unit patient-days on which the magnesium protocol was active, not as a measure of adherence (because there was no restriction on discontinuing the protocol) but to give a sense of the proportion of critical illness time covered by the intervention.

#### Item 19b. Description of how adherence to the intervention will be presented

Adherence will be reported as a proportion. The denominator will include all magnesium levels resulted while the magnesium protocol is active. The numerator will include all such magnesium levels where the recommended action was consistent with the assigned protocol, as defined in Item 19a. We will also report the adherence proportion according to treatment assignment and whether magnesium supplementation was recommended by protocol, and the number and proportion of patients overall and by arm for whom adherence was perfect.

As an additional adherence sensitivity analysis, we will calculate adherence using Bayesian logistic regression with a fixed covariate for treatment assignment and a random intercept for each patient (to account for multiple measurements per patient). This will be used to compare adherence between the treatment strategies.

#### Item 19c. Definition of protocol deviations for the trial

In addition to the adherence definition above, we will define administration of magnesium not indicated by the protocol, while the protocol was ordered, as a protocol deviation. We will not assess the rationale for administrations of magnesium that deviate from protocol, nor the rationale for not administering magnesium that was mandated by protocol, because we anticipate that the data regarding rationale in the EHR will be incomplete.

#### Item 19d. Description of which protocol deviations will be summarized

We will summarize the number and proportion of patients who had a protocol deviation for each of the two possible types of deviations: magnesium administered when the most recent magnesium level would not mandate protocolized supplementation, and magnesium not administered within 8 hours of a magnesium level which would mandate protocolized supplementation.

#### Item 20. Definition of analysis populations, e.g., intention-to-treat, per-protocol, complete case, safety

The primary analysis population will be an intention-to-treat population defined by the assigned protocol. Because the primary outcome is coded into the electronic health record, we do not anticipate missing outcome data, and the primary analysis population will be a complete case population with respect to the primary outcome.

### Section 5: Trial Population

#### Item 21. Reporting of screening data (if collected) to describe representativeness of the trial sample

We will report the proportion of patients admitted to the intensive care unit for whom a magnesium protocol was ordered.

#### Item 22. Summary of eligibility criteria

Inclusion criteria:

1) Adult (age 16 years or more)
2) Order placed for admission to a medical-surgical intensive care unit (ICU)
3) Order placed for the magnesium replacement protocol.

Exclusion criteria:

1) Current diagnosis of eclampsia or pre-eclampsia
2) Ongoing concern for sustained ventricular tachycardia
3) Active neuromuscular junction disease (e.g. myasthenia gravis)
4) Prior enrollment in or withdrawal from the MAGNOLIA trial
5) Enrollment in a separate trial that prevents administration of the MAGNOLIA intervention (for example, a trial of magnesium supplementation vs no supplementation)

#### Item 23. Information to be included in the CONSORT flow diagram

We will report the number of patients who met inclusion criteria for the study, the number who were deemed ineligible (and summarized reasons for ineligibility), the number assigned to each treatment, the number who were withdrawn by patient or clinician request, and the number for whom the primary outcome is available.

#### Item 24a. Level of withdrawal, e.g., from intervention and/or from follow-up

All patients who withdraw from the trial do so after at least some duration of intervention. Withdrawals initiated by the patient or their substitute decision maker will have their data removed from the trial, while withdrawals initiated by the physician team will still contribute data to the analysis under their assigned treatment category.

#### Item 24b. Timing of withdrawal / lost-to-follow-up data

We will report the number of withdrawals that happen within the 30-day post-enrollment period when the intervention is active, and the number of withdrawals that happen after the 30-day post-enrollment period.

#### Item 24c. Reasons and details of how withdrawal / lost-to-follow-up data will be presented

We will report the number of withdrawals that were initiated by the patient or their substitute decision-maker, and the number that were initiated by the most responsible physician. Where possible, we will summarize the reasons for withdrawal in both groups.

#### Item 25a. List of baseline characteristics to be summarized

Patients will be described with respect to age, sex, and hospital length of stay prior to enrollment; comorbidities of atrial fibrillation or flutter, diabetes, ventricular tachycardia, end-stage renal disease, and congestive heart failure; primary diagnosis by category, Sequential Organ Failure Assessment (SOFA) score, receipt of vasopressors, receipt of invasive ventilation and most recent documented cardiac rhythm at time of enrollment; most recent creatinine, magnesium, and lactate available at time of enrollment. We will describe both the overall enrolled trial population and the population assigned to each treatment.

SOFA score will be calculated by imputing zero for missing components, consistent with the original design of the score.[16] For patients without an arterial partial pressure of oxygen, we will use a previously described conversion to the saturation-to-arterial partial pressure of oxygen ratio (SOFA 1 / 2 / 3 / 4 at Spo2/FiO2 < 512 /  357 / 214 89).[17] Where there is no inspired oxygen fraction charted, we will convert low-flow oxygen to an inspired oxygen fraction using the equation 0.21 + 3*(oxygen flow in liters per minute).[18]

#### Item 25b. Details of how baseline characteristics will be descriptively summarized

Categorical data will be summarised by numbers and percentages. Continuous data will be summarised by mean, SD and range if data are normal and median, interquartile range (IQR) if data are skewed. Tests of statistical significance will not be undertaken for baseline characteristics, but the clinical importance of any imbalance will be noted.

### Section 6: Analysis

### Item 26a. Specification of outcomes and timings

Our outcomes include primary and secondary outcomes, including some Desirability Of Outcome Ranking (DOOR) outcomes.[19]

| Type | Definition |
|---|---|
| Primary | Days alive and free of atrial fibrillation or flutter - an ordinal composite combining hospital mortality at day 30 (scored as -1) and days free of atrial fibrillation or flutter[20] in ICU at day 30 |
| Secondary: evaluated at 30 days | Hospital mortality<br>Days alive and free of ICU<br>Days alive and free of organ support (vasopressors, invasive ventilation, or new renal replacement therapy)<br>Days alive and free of invasive ventilation<br>Days alive and free of vasopressors<br>Sustained ventricular tachycardia or ventricular fibrillation in ICU<br>Receipt of an intravenous antiarrhythmic*<br>Magnesium and potassium levels while in ICU (mean, standard deviation, proportion below the lower limit of normal; all reported overall and by study day)<br>Magnesium administrations while in ICU (number, route, dose, amount)<br>Daily fluid balance while in ICU<br>Cost of magnesium administrations while in ICU<br>Carbon footprint of magnesium administrations while in ICU |
| Secondary: DOOR, evaluated at 30 days | DOOR 1: Death, ventricular tachycardia or ventricular fibrillation, atrial fibrillation or flutter, none of the above<br>DOOR 2: Death, persistent organ dysfunction (ongoing vasopressors, invasive ventilation, or continuous renal replacement therapy) in ICU, ongoing hospital admission, discharge to location other than home prior to day 30, discharge to home prior to day 30. |
| Secondary: 60 or 90 days | Hospital mortality at day 60<br>Hospital mortality at day 90<br>Hospital length of stay at day 90<br>Discharge destination from last observed hospitalization up to day 90 |

* Intravenous antiarrhythmic defined as amiodarone, metoprolol, esmolol, diltiazem, procainamide, lidocaine, flecainide, adenosine, or digoxin

### Item 26b and 26c. Specific measurement and units, with transformations or calculations

| Outcome | Units and measurement details |
|---|---|
| Primary outcome: days alive and free of atrial fibrillation or flutter (in the ICU) at 30 days | Ordinal: scale from -1 (death) to 30 (survival with no atrial fibrillation or flutter noted during ICU stay) |
| Days alive and free of ICU | Ordinal: Ranging from 0 to 30 |

| | |
|---|---|
| Days alive and free of organ support (vasopressors, invasive ventilation, and renal replacement therapy)<br>Days alive and free of invasive ventilation<br>Days alive and free of vasopressors | |
| Sustained ventricular tachycardia or ventricular fibrillation in ICU<br>Receipt of an intravenous antiarrhythmic<br>Hospital mortality at day 30<br>Hospital mortality at day 60<br>Hospital mortality at day 90<br>Hospital length of stay at day 90 | Binary: yes/no |
| Magnesium and potassium levels while in ICU | Continuous: mmol/L |
| Magnesium administrations while in ICU (number, route, dose) | Number – count per day; route – enteral or intravenous; dose – mmol per day |
| Daily net fluid balance while in ICU | Continuous: mL/day |
| Discharge destination from last observed hospitalization up to day 90 | Ordinal: Home without Support / Home with Support* / Other institution or retirement home / Still in hospital / Death |
| Costs | Continuous: 2026 Canadian dollars (CAD) |
| Carbon footprint | Continuous: kilograms of carbon dioxide equivalent ($kgCO_2e$) |

* Support defined as home nursing or personal support worker care

### Item 27a. What analysis method will be used and how treatment effects will be presented

*Primary outcome:* The number and percentage of deaths, and patients with atrial fibrillation or flutter by 30 days after enrollment, will be reported for each treatment group. We will report the median number of days with atrial fibrillation or flutter in the ICU among patients who survived and had at least one day of atrial fibrillation or flutter in the ICU. We will generate a horizontal stacked bar chart to show the distribution of the primary outcome in each treatment group. In the appendix, we will include a histogram of the number of patients (pooled across arms) with each of the 32 possible composite primary outcome values.

*Primary analysis:* The primary effect estimate will be the odds ratio of a more favourable outcome with respect to the ordinal composite primary outcome of mortality and days free of atrial fibrillation or flutter in the ICU at 30 days, comparing higher target to lower target. The primary effect will be calculated using a Bayesian proportional odds model with a normally-distributed random effect by hospital site. The treatment effect coefficient will be a weakly skeptical prior distribution (normal(0,1) in log-odds space), and the prior distribution for the standard deviation of the site-level random effect will be a half-normal distribution with variance of 1.[21,22] Ordinal outcome intercept priors will be constrained to be ordered and modeled using Student-t distributions with 3 degrees of freedom, location 0 and scale 2.5.[23] We will report the posterior mean and 95% credible interval for the odds ratio. See Figure 1 for equations.

The primary analysis will be fit using the No-U-Turn Sampler, an adaptive variant of Hamiltonian Monte Carlo, implemented in Stan and accessed through the R package brms. We will run 4 chains of 2,000 iterations each (1,000 warm-up) and declare adequate model convergence if R-hat values are < 1.01, bulk and tail effective sample sizes are > 400, and there are no divergent transitions.[24]

If the primary analysis does not meet those convergence criteria, we will collapse the 32-level outcome distribution into 5 levels as follows: from best to worst: 30 (survival with no atrial fibrillation or flutter in ICU), 29 (survival with 1 day with atrial fibrillation or flutter in ICU), 27-28 (survival with 2-3 days with atrial fibrillation or flutter in ICU), 0-26 (survival with 4 or more days with atrial fibrillation or flutter in ICU), -1 (death). Using this simplified outcome, we will repeat the primary analysis with the same prior distributions specified above.

*Secondary outcomes:* We will report the number and percentage of binary outcomes, the median and interquartile range of integer outcomes, and the mean with standard deviation of continuous outcomes according to treatment assignment.

*Secondary analyses:* Treatment effects for secondary outcomes will be drawn from Bayesian analyses using regression with the same prior distributions and site-level random effects as above. The regression will vary according to the nature of the outcome: logistic regression for binary outcomes, proportional odds regression for ordinal outcomes, and linear regression for continuous outcomes; similarly, the treatment effect reported will be an odds ratio (logistic regression or proportional odds regression) or an absolute difference (linear regression). For all treatment effects we will report the posterior mean and 95% credible interval.

The primary and secondary analyses that yield odds ratios will be summarized in a forest plot showing the posterior mean and 95% credible intervals.

### Figure 1: Equations for primary analysis and proportional odds assumption check

#### A. Primary analysis: Bayesian proportional odds model

$$\text{logit}\left[\Pr\left(Y_{ij} \geq k\right)\right] = \alpha_k + \beta x_{ij} + u_j, \qquad k = 2, \ldots, 32$$

*$Y_{ij}$: ordinal composite outcome at 30 days for patient $i$ at site $j$, coded worst to best: $Y_{ij} = 1$ for death, otherwise $Y_{ij} = 2 + D_{ij}$, where $D_{ij} \in \{0, \ldots, 30\}$ is days alive and free of AF/flutter in the ICU (32 levels). $x_{ij} = 1$ for higher target and 0 for lower target. $\alpha_k$: ordered intercepts. $u_j$: site-level random effect.*

$$u_j \sim N\left(0, \sigma_u^2\right), \qquad \beta \sim N(0, 1), \qquad \alpha_k \sim t_3(0,\, 2.5), \qquad \sigma_u \sim \text{Half-Normal}(0, 1)$$

*Prior distributions: weakly sceptical normal prior on the treatment effect (log-odds scale); Student-t prior with 3 degrees of freedom, location 0 and scale 2.5 on the intercepts, constrained to be ordered; and half-normal prior on the between-site standard deviation.*

$$\text{OR} = e^{\beta}$$

*Odds of a more favourable outcome with higher versus lower target, reported as the posterior mean and 95% credible interval.*

#### B. Proportional odds check: partial proportional odds model

$$\text{logit}\left[\Pr\left(Y_{ij} \geq k\right)\right] = \alpha_k + \beta x_{ij} + \gamma\, x_{ij}\, I(k = 2) + u_j, \qquad \gamma \sim N(0, 1)$$

*$I(k = 2) = 1$ for the survival dichotomy (alive vs. dead) and 0 otherwise, so $\gamma$ allows the treatment effect on mortality to differ from its effect on days free of AF/flutter. All other terms and priors are as in panel A.*

$$\text{OR}_{\text{AF}} = e^{\beta}, \qquad \text{OR}_{\text{mort}} = e^{\beta+\gamma}$$

*$\text{OR}_{\text{AF}}$: odds ratio across the days-free-of-AF/flutter dichotomies. $\text{OR}_{\text{mort}}$: odds ratio for survival.*

$$\max\left[\Pr\left(\gamma > 0 \mid \text{data}\right), \Pr\left(\gamma < 0 \mid \text{data}\right)\right] > 0.90 \quad \text{and} \quad \text{sign}\left(\hat{\beta} + \hat{\gamma}\right) \neq \text{sign}\left(\hat{\beta}\right)$$

*The proportional odds assumption is considered violated if both conditions hold: the mortality and AF/flutter effects differ with >90% posterior probability, and their posterior means (hats) fall on opposite sides of an odds ratio of 1.*

### Item 27b. Any adjustment for covariates

The primary and secondary analyses will include site-level random effects, with no adjustment for fixed covariates.

### Item 27c. Methods used to check assumptions for statistical methods

The proportional odds assumption is the most relevant assumption for our primary analysis. The odds ratio calculated by a proportional odds model can be seen as the average odds ratio across all dichotomies of the ordinal outcome. Because our primary outcome is a composite of mortality and days alive and free of atrial fibrillation in the ICU, it is possible that the intervention could affect mortality and days alive and free of atrial fibrillation in opposing ways (i.e. reduce mortality but increase days with atrial fibrillation, or vice versa). It is not plausible that the intervention would have effects in different directions across the "days free of atrial fibrillation or flutter in the ICU" portion of the outcome. Therefore, we will focus our efforts on checking that the proportional odds model with respect to mortality versus days alive and free of atrial fibrillation or flutter in the ICU is met.

To do so, we will fit a partial proportional odds model, allowing the intervention's effect on mortality to differ from its effect on days alive and free of atrial fibrillation or flutter. If the odds ratio for mortality has a greater than 90% probability of differing from the odds ratio across the days alive and free of atrial fibrillation or flutter, and the odds ratio for mortality lies on the opposite side of equivalence from the point estimate of the odds ratio for days alive and free of atrial fibrillation or flutter, we will consider the proportional odds assumption to be violated. See Figure 1 for equations.

**Item 27d. Details of alternative methods if distributional assumptions do not hold**

If the procedure in 27c does not support the proportional odds assumption, we will report the odds ratios for mortality and days free of atrial fibrillation from the partial proportional odds model separately.

**Item 27e. Any planned sensitivity analyses for each outcome, where applicable**

We will perform the primary analysis on the subgroup of patients who have no evidence of atrial fibrillation at enrolment, to provide an estimate of the extent to which higher compared to lower targets for protocolized magnesium supplementation affect the incidence of new-onset atrial fibrillation. Note that this differs subtly from a subgroup analysis, where the goal is to evaluate whether the intervention effect differs according to subgroup membership. We will also perform a subgroup analysis to assess whether the effect of the intervention differs according to baseline atrial fibrillation status. The difference is that the sensitivity analysis will have slightly less uncertainty (slightly more statistical power) than the treatment estimate in patients without atrial fibrillation at baseline that is derived from the subgroup analysis.

**Item 27f. Any planned subgroup analyses for each outcome, including how subgroups are defined**

We will perform pre-specified subgroup analyses modeling the interaction between subgroup membership and treatment effect with Bayesian regression. Pre-specified subgroups will include: male vs female sex (hypothesis: higher target benefits those with female sex), age quintiles (hypothesis: higher target benefits older patients), atrial fibrillation at baseline or not (hypothesis: higher target benefits those without atrial fibrillation at baseline), primary diagnosis of acute coronary syndrome or not (hypothesis: higher target benefits those with acute coronary syndrome), receiving vasopressors at baseline or not (hypothesis: higher target benefits those receiving vasopressors at baseline), invasively ventilated at baseline or not (hypothesis: no difference), end-stage renal disease at baseline or not (hypothesis: higher target benefits those without end-stage renal disease), and primary diagnosis of sepsis or not (hypothesis: higher target benefits those with sepsis). Credibility of subgroup effects will be assessed using the Instrument to assess the Credibility of Effect Modification Analyses (ICEMAN).[25]

**Item 28. Reporting and assumptions / statistical methods to handle missing data**

We will not impute missing outcome data. Patients with missing outcome data will be removed from the analysis concerning that outcome. Due to the nature of our outcomes and the trial's integration with the electronic health record, we anticipate < 1% missing data.

**Item 29. Details of any additional statistical analyses required**

We also plan a heterogeneity of treatment effect analysis whose full statistical analysis plan will be described separately and posted on a publicly-available repository in advance of conducting that analysis.

Similarly, we plan to investigate treatment effect and outcomes in more detail in the following subsets of patients:

- Patients with cardiogenic shock
- Patients with chronic obstructive pulmonary disease exacerbations
- Patients with sepsis and septic shock
- Patients who received vascular surgery
- Patients who received thoracic surgery

We also plan to perform a secondary analysis of pain scores and analgesia according to assigned target, and to analyze and report on the trial process, including time taken to screen each patient and time of day of enrolments.

**Item 30. Sufficient detail on summarizing safety data: severity, expectedness, causality; how adverse events are coded/categorized; how adverse event data will be analysed (e.g., grade 3/4 only, incidence case analysis, intervention-emergent analysis)**

Our safety data for this pragmatic trial relies on surveillance of incidents reported via the hospital quality improvement and incident reporting system. We will report the number of incidents pertaining to magnesium administration that occurred in patients during their first 30 days of enrollment in the MAGNOLIA trial. We will also report the number of occasions on which a magnesium level > 2 mmol/L was recorded, and in how many patients this occurred.

**Item 31. Details of statistical packages to be used to carry out analyses**

We will use R as the statistical software for analysis, including the tidyverse package for data processing and presentation, and the brms package for Bayesian analyses, which uses Stan as the underlying sampler.[23,26–28]

**Item 32a. References for nonstandard statistical methods**

- Proportional odds modeling[29]
- Partial proportional odds modeling[30,31]

#### Item 32b. Data Management Plan

Data management is described in the trial protocol.

#### Item 32c. Reference to the Trial Master File and Statistical Master File

The statistical analysis code will be pre-registered on a publicly available online platform (preprint or Open Science Framework or similar) prior to the completion of follow-up for the primary outcome for the last enrolled patient.

#### Item 32d. Reference to other standard operating procedures or documents to be adhered to

This protocol was written using the guidance of Gamble et. al. and their associated statistical analysis plan checklist.[15]
